# The effect of special educational assistance in early childhood education and care on psycho-social difficulties in elementary school children

**DOI:** 10.1101/2021.02.18.21251836

**Authors:** Guido Biele, Ratib Lekhal, Kristin R. Overgaard, Mari Vaage Wang, Ragnhild Eek Brandlistuen, Svein Friis, Paal Zeiner

## Abstract

Three to seven percent of preschoolers have developmental problems or child psychiatric disorders. Randomized controlled trials (RCTs) indicate that interventions in early childhood education and care improve long-term outcomes of children from disadvantaged backgrounds. It is unknown if effects generalize beyond the well-structured context of RCTs and to children who may not have a disadvantaged background but have developmental problems or psychiatric disorders. We use data from the population-based Norwegian Mother, Father and Child Cohort Study, recruiting pregnant women from 1999 to 2008, with child follow-up from ages 6, 18, and 36 months to ages 5, 7, and 8 years. This sub-study included 2499 children with developmental problems or psychiatric disorders at age five. We investigate the effects of special educational assistance at age five on mother-reported internalizing, externalizing, and communication problems at age eight. We analyze bias due to treatment by indication with directed acyclic graphs, adjust for treatment predictors to reduce bias, and estimate effects in different patient groups and outcome domains with a hierarchical Bayesian model. In the adjusted analysis, preschoolers with special educational assistance had on average by 0.1 (0.03-0.16) standardized mean deviation weaker psycho-social difficulties in elementary school. Mean effect sizes varied between groups and outcomes. We estimate positive effects of educational assistance during the transition from preschool to the school years. It should therefore be considered as an intervention for preschoolers with developmental or behavior problems. More research with improved measurements of treatment and outcomes is needed to identify success factors for their implementation.

---

Between three and seven percent of preschoolers have developmental problems or child psychiatric disorders [1, 2], which are an important risk factor for mental disorders in adulthood [3]. Efforts to promote healthy growth and development in children who struggle in the early years can accordingly improve children’s long-term life opportunities [4]. This effect seems to decrease as children grow older. Therefore, investing resources later, at the age of school entry or beyond, may show less of an effect [5].

Interventions in early childhood are often described as an effective method to improve the long-term outcomes of children from disadvantaged backgrounds [6] or those with specific developmental or behavioral problems like attention deficit hyperactivity disorder, autism, or behavior or language problems [7]. Interventions in early childhood education and care (ECEC) can be especially effective because in contrast to parental training programs, their implementation relies less on parents’ abilities or motivation, and on average 93% of three to five year old children in Organisation for Economic Co-operation and Development (OECD) countries are enrolled in ECEC [8]. Randomized controlled trials (RCTs) reported clear effects of early interventions in ECEC for a horizon of up to nine months, for instance for language problems [9], children with ADHD or autism [10], and for teacher classroom management programs [11].

However, the effect sizes of such interventions are not generally large, and less is known about their effect when interventions are provided outside the well-structured context of RCTs. Even though RCTs are, due to their interval validity, the gold standard for estimating treatment effects, differences between study sample and target population and differences in treatment-implementation between study and regular care contexts, make a generalization of findings from RCT samples to populations of interest difficult [12–15]. Since RCTs often take place in a controlled setting, it may be difficult to replicate the results in other, less rigid settings. For instance, field professionals in ECEC institutions will draw on a much wider range of sources than formal experimental evidence in order inform their actions. Thus, while evidence from RCTs is encouraging, it remains unclear how it generalizes to interventions in ECEC provided in regular care.

Only a handful of studies examined the effects of special educational assistance (SEA) interventions in ECEC when they are implemented outside of RCTs. These studies used propensity scores to deal with the problematic internal validity in observational studies—due to treatment by indication—and found that children who received SEA in ECEC showed the same or worse outcomes compared children who did not receive SEA [16, 17].

The Norwegian ECEC-system facilitates the investigation of SEA, because children who cannot fully benefit from standard education and care have the right to receive free SEA. Similar to other OECD countries [2], around 4.5% of preschoolers in Norwegian ECEC have impaired functioning, the most common impairment being language and communication difficulties, followed by psycho-social difficulties and behavior problems [18]. Around 2.6% of preschoolers receive SEA, which is provided for several hours per week for individual children [18]. After stimulation of language development, social- and behavior-training are the most frequent types of SEA provided. To date, no study has–to the best of our knowledge–examined the effect of SEA in ECEC on children’s psycho-social difficulties. Related studies on SEA in Norwegian schools report that students who received it have similar or slightly worse scholastic outcomes compared to those who did not receive it [19, 20, see also 21].

In sum, the few studies examining effects of SEA in ECEC outside the context of RCTs reported small negative, to no effects of SEA. Moreover, most studies focused on educational outcomes, such that the effect of SEA on the development of psycho-social difficulties remains largely unclear. Hence, this large-scale prospective cohort study adds to the existing literature by investigating how SEA in ECEC provided outside RCTs affects the psycho-social development of children with developmental or behavioral problems.

## Methods

### Participants

The sample is a sub-sample of the Norwegian Mother, Father and Child Cohort Study (MoBa), a prospective population-based pregnancy cohort study conducted by the Norwegian Institute of Public Health [22, 23]. Participating mothers from all over Norway were recruited during routine ultrasound assessment in week 17 or 18 of their pregnancy in the period from 1999 to 2009. 41% of the invited women consented to participate. MoBa participants received questionnaires in gestational week 17 or 18, week 22 and week 30, at child’s age 6 and 18 months, 3, 5, and 8 years and onward. The study is still ongoing. The reported analyses also use information from the Medical Birth Registry of Norway [24]. Figure 1 shows the inclusion-flowchart.

**Figure 1.**
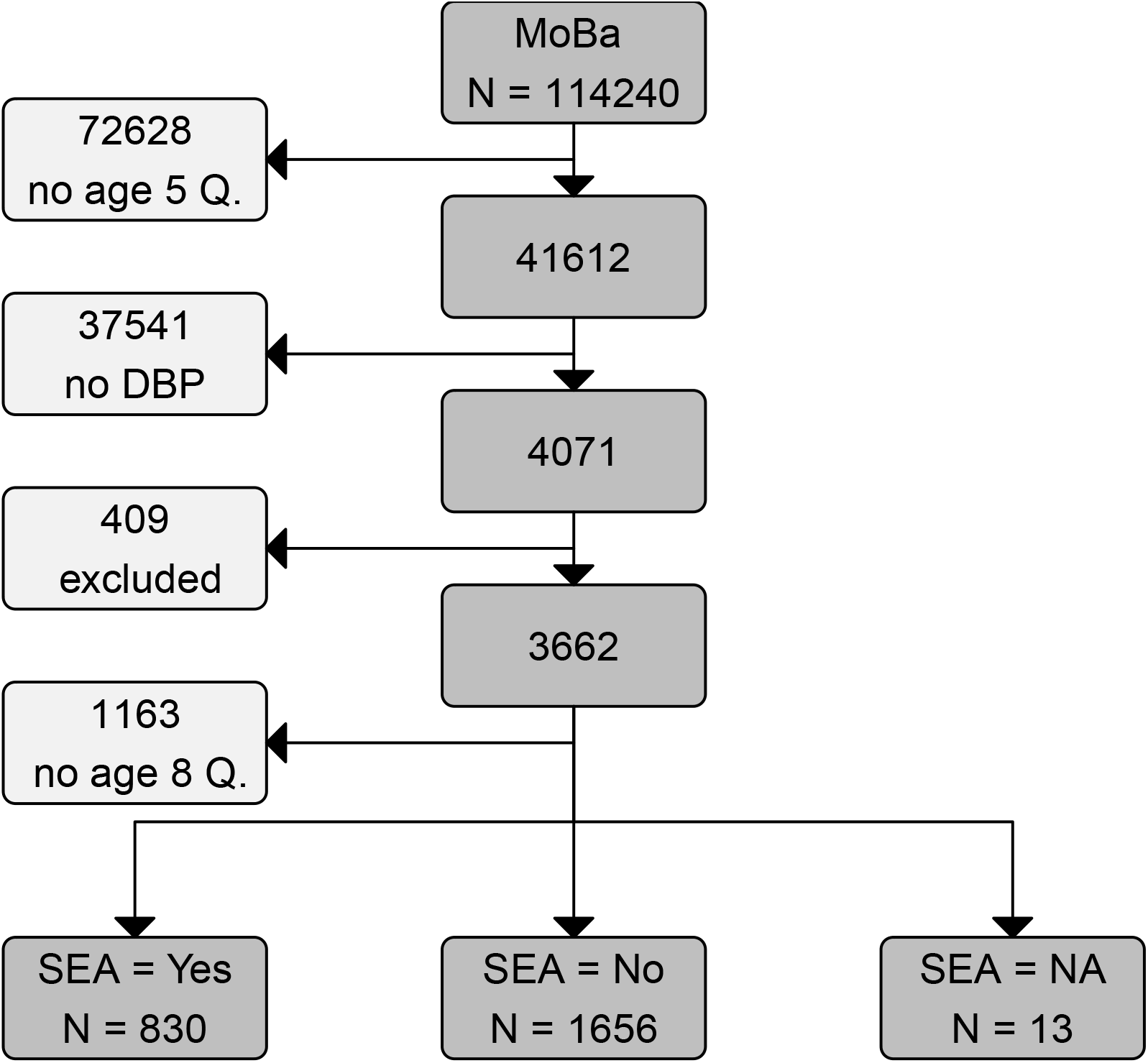
Inclusion flow chart. Age 5 and 8 Q are MoBa questionnaires sent out at child age five and eight years. Children in the study sample have a parent-reported developmental or behavior problem (DBP) at age five. Children with epilepsy, cerebral palsy, chromosomal defects, severe developmental delay, or hearing loss were excluded from this study. SEA = special educational assistance in early childhood education and care (ECEC).

The study sample is comprised of children whose mothers indicated developmental or behavior problems in MoBa’s age five years questionnaire, and for whom information about outcomes in the age eight years questionnaire are available. This study focuses on children with one or more of the following developmental or behavioral problems: Attention deficit hyperactivity disorder, language development, oppositional defiant or conduct disorder, autism spectrum disorder, and learning disabilities.

### Materials

The current study used rating scales from MoBa questionnaires sent out at child ages five, and eight years. Exposure and inclusion criteria were based on responses in the five year questionnaire, whereas outcome measures were taken from the eight year questionnaire. The first, 1.5 and three year MoBa questionnaires and the Medical Birth Registry of Norway provided covariates.

#### Exposure

To measure the provision of SEA, we relied on following question: “Does your child receive, or has received any extra resources in the kindergarten?” If mothers responded “Yes” to this question, they were additionally asked about the number of hours per week. SEA is provided to individual children, both inside and outside the context of regular preschool activities.

#### Outcome variables

Outcome variables (*PSD*_8_ in Figure 2) were *sum scores* from different scales about psycho-social difficulties. Outcome dimensions were attentional, hyperactivity/impulsivity, and behavioral (ODD or CD) problems measured with the Parent Rating Scale for Disruptive Behavior Disorders (RS-DBD, [25]), emotional problems measured with the Short Mood and Feelings Questionnaire (SMFQ, [26]) and the Screen for Child Anxiety Related Disorders (SCARED, [27]), and communication problems measured with the Children’s Communication Checklist-2 (CCC-2, [28]).

**Figure 2.**
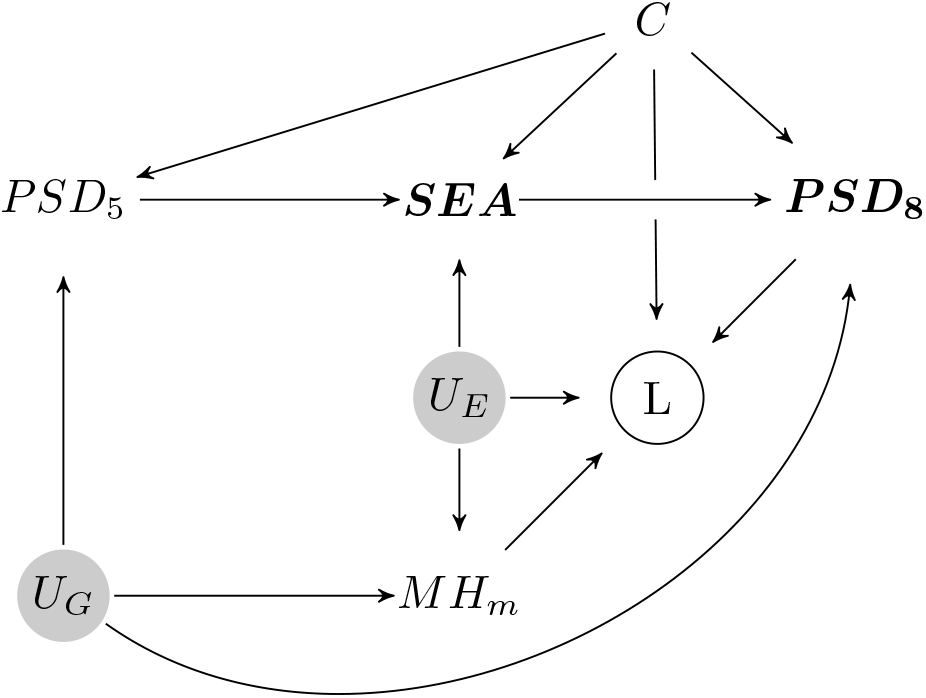
Directed Acyclic Graph of the hypothesized causal relationships between special educational assistance (*SEA*) psycho-social difficulties (*PSD*_*5*_, *PSD*_*8*_), loss to follow up (*L*), maternal mental health (*MH*_*m*_), unobserved environmental and genetic causes (*U*_*E*_,*U*_*G*_) and additional confounders (*C*, i.e. contact with mental health services, maternal education, birth order, birth month, preterm birth). Current and prior psycho-social difficulties *PSD*_*5*_ are confounders causing bias due to treatment by indication and can be controlled through adjustment. Because maternal mental health (*MH*_*m*_) predicts loss to follow up (*L*), which is a collider on a backdoor path between *SEA* and *PSD*_*8*_, loss to follow up has to be controlled through inverse probability weighting.

#### Adjustment variables

Adjustment variables and those to control for loss to follow up were chosen based on the directed acyclic graph (DAG) shown in figure Figure 2. One important set of confounders includes children’s psycho-social difficulties at baseline, because these can be seen as causes of treatment and are related to later psycho-social difficulties. A number of scales in MoBa assessed psycho-social difficulties at age five and served as baseline measures (*PSD*_5_ in Figure 2). These included the Conners’ Parent Rating Scale-Revised, Short Form (CPRS-R (S), [29]), Child Behavior Checklist (CBCL, [30]), the Ages and Stages Questionnaire (ASQ, [31]), and the Children’s Communication Checklist-2.

Additional variables used for adjustment or prediction of loss to follow-up included maternal age, education, and mental health (ADHD symptoms measured with the Adult ADHD Self-Report Scale [32] at child age three and depressive symptoms measured with the SCL-5 [33] at child age five, parity, preterm birth, birth-month, hours special education per week, number of developmental of behavior problems, and contact with rehabilitation services, Child and Adolescent Psychiatric Units, or Educational and Psychological Counseling Service at child age five years.

### Classification into groups with different developmental or behavioral problems

To classify if and in which area a child had developmental or behavioral problem (DBP), we used MoBa questions about mental health problems at age five. Mothers were asked if their child “suffered, or is currently suffering from any of the following long-term illnesses or health problems.” In addition, mothers were asked if they had been in contact with a Child and Adolescent Psychiatric Unit or the Educational Psychology Counseling Services and if the health problem was confirmed by a professional. Only children for whom mothers reported a health problem *and* who indicated that the problem was evaluated by a mental health professional were included in the sample.

Disorders or health problems for which MoBa’s age 5 questionnaire has questions included Epilepsy, Cerebral Palsy, impaired hearing, which were excluded from the current analysis, together with children for whom mothers indicated a chromosomal defect. MoBa also asked mothers about autism spectrum disorders (ASD), hyperactivity and attention problems (ADHD), language difficulties (Lang), and behavioral problems (Beh). Additional questions about learning disabilities (LD) were also used to identify cases of interest for this study. Each child was classified in one of the following DBP groups: 1. ASD, 2. LD, 3. ADHD & Beh & Lang, 4. ADHD & Beh, 5. ADHD & Lang, 6. ADHD, 7. Lang, 8. Beh. For some children, mothers indicated multiple DBP, in which case the child was assigned to the first group it fell into. If, for example, a mother indicated ASD, ADHD, and language problems, the child was assigned to the ASD group (details in supplementary materials and Table S1). The rational underlying this classification scheme was to use existing psychiatric diagnoses, and to classify children according to their most impairing problem because these have typically more severe and persistent effects psycho-social on psycho-social development.

### Data analysis

All analyses were performed using R [34]. The Bayesian hierarchical regression model was implemented with the brms package [35]. The analyes are described in more detail in the supplementary material, and analysis scripts are available at https://github.com/gbiele/SPS358.

#### Bias from treatment by indication and loss to follow up

Estimation of treatment effects from observational data is difficult because treatment is not assigned randomly. Instead, individuals with more psycho-social difficulties at age five, who are also more likely to have psycho-social difficulties in the future, more likely receive treatment (treatment by indication). In addition, loss to follow up makes estimation of treatment effects difficult. Therefore, we used a directed acyclic graph [DAG, 36, see Figure 2] to explicate the assumed causal structure and to determine with which approach to deal with potential biases. Given this structural model, inverse probability of continued participation weighting was needed to reduce bias from loss to follow up [37], whereas adjustment for predictors of SEA was sufficient to control bias from treatment by indication. This means that we effectively estimated the effect of SEA on the change of psycho-social difficulties from preschool to elementary school.

#### Estimation of the treatment effects

We used a Bayesian adjusted and weighted hierarchical ordinal regression to estimate effects of SEA [35, 38, 39]. A hierarchical regression induces partial pooling (shrinkage) of estimates, which reduces the variance of estimates [40] and controls the multiple comparison problem [41]. Importantly, when anlysing related patient groups hierarchical regression results in more accurate association estimates then independent analysis of these groups [40]. We used an ordinal regression model, because the estimation of latent, normally distributed traits that underlie the rating-scale responses facilitates the presentation of results in terms of standardized mean differences (SMD). The reported results were obtained by pooling over the independent analyses of the 50 imputed data sets [42]. Consistent with recent recommendations to focus on estimation of effect sizes instead of significance testing [43, 44] we generally report means and the 90% credible intervals.

## Results

The study sample includes 2499 participants (c.f., Figure 1). Thirty-three percent of the children in the sample received SEA. Table 1 describes the study sample. Figures S4 and S5 show that children with more severe problems (e.g. ASD) were more likely to receive SEA and also received SEA from better educated personnel.

**Table 1.**
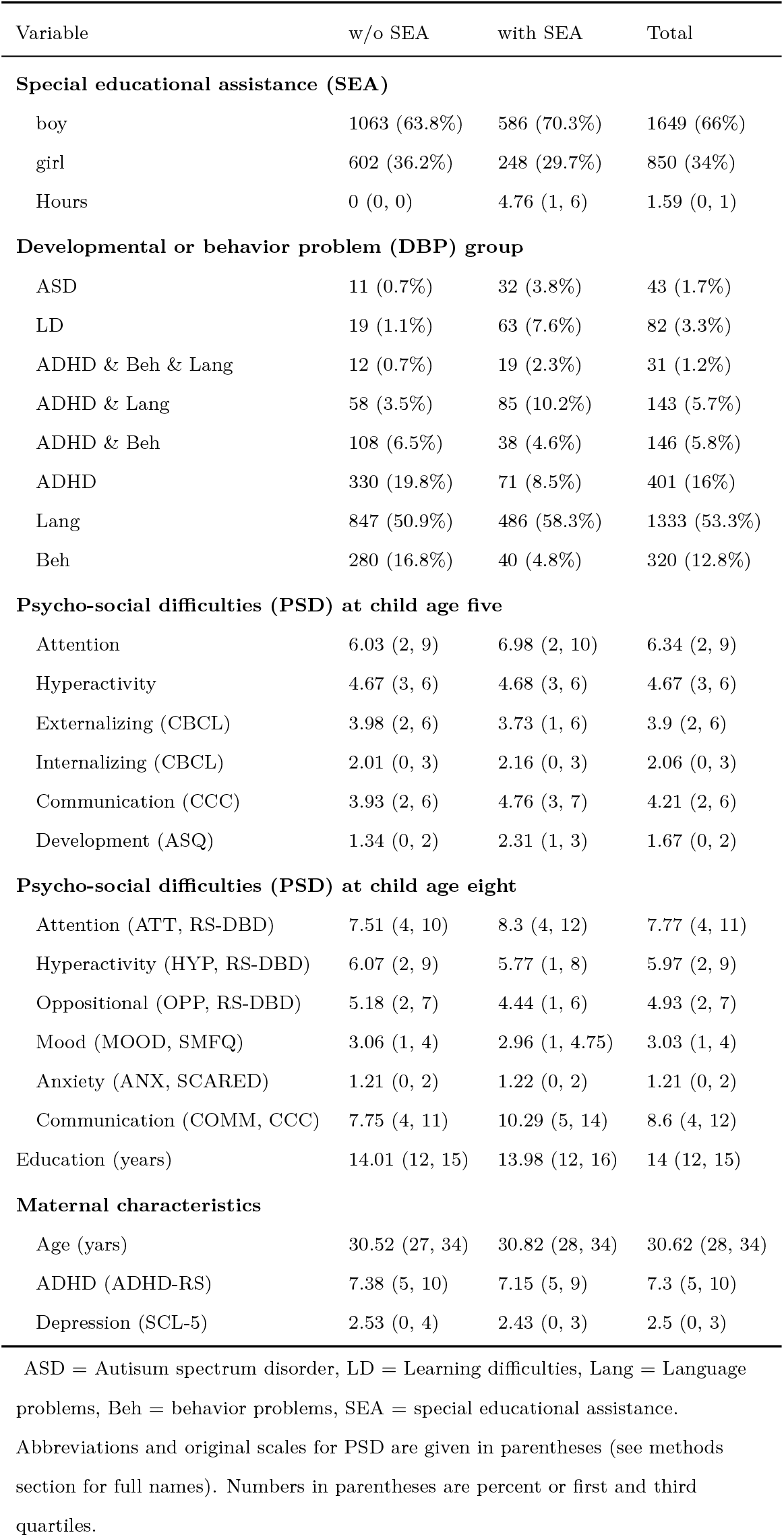
Study sample

| Variable | w/o SEA | with SEA | Total |
| --- | --- | --- | --- |
| **Special educational assistance (SEA)** |  |  |  |
| boy | 1063 (63.8%) | 586 (70.3%) | 1649 (66%) |
| girl | 602 (36.2%) | 248 (29.7%) | 850 (34%) |
| Hours | 0 (0, 0) | 4.76 (1, 6) | 1.59 (0, 1) |
| **Developmental or behavior problem (DBP) group** |  |  |  |
| ASD | 11 (0.7%) | 32 (3.8%) | 43 (1.7%) |
| LD | 19 (1.1%) | 63 (7.6%) | 82 (3.3%) |
| ADHD & Beh & Lang | 12 (0.7%) | 19 (2.3%) | 31 (1.2%) |
| ADHD & Lang | 58 (3.5%) | 85 (10.2%) | 143 (5.7%) |
| ADHD & Beh | 108 (6.5%) | 38 (4.6%) | 146 (5.8%) |
| ADHD | 330 (19.8%) | 71 (8.5%) | 401 (16%) |
| Lang | 847 (50.9%) | 486 (58.3%) | 1333 (53.3%) |
| Beh | 280 (16.8%) | 40 (4.8%) | 320 (12.8%) |
| **Psycho-social difficulties (PSD) at child age five** |  |  |  |
| Attention | 6.03 (2, 9) | 6.98 (2, 10) | 6.34 (2, 9) |
| Hyperactivity | 4.67 (3, 6) | 4.68 (3, 6) | 4.67 (3, 6) |
| Externalizing (CBCL) | 3.98 (2, 6) | 3.73 (1, 6) | 3.9 (2, 6) |
| Internalizing (CBCL) | 2.01 (0, 3) | 2.16 (0, 3) | 2.06 (0, 3) |
| Communication (CCC) | 3.93 (2, 6) | 4.76 (3, 7) | 4.21 (2, 6) |
| Development (ASQ) | 1.34 (0, 2) | 2.31 (1, 3) | 1.67 (0, 2) |
| **Psycho-social difficulties (PSD) at child age eight** |  |  |  |
| Attention (ATT, RS-DBD) | 7.51 (4, 10) | 8.3 (4, 12) | 7.77 (4, 11) |
| Hyperactivity (HYP, RS-DBD) | 6.07 (2, 9) | 5.77 (1, 8) | 5.97 (2, 9) |
| Oppositional (OPP, RS-DBD) | 5.18 (2, 7) | 4.44 (1, 6) | 4.93 (2, 7) |
| Mood (MOOD, SMFQ) | 3.06 (1, 4) | 2.96 (1, 4.75) | 3.03 (1, 4) |
| Anxiety (ANX, SCARED) | 1.21 (0, 2) | 1.22 (0, 2) | 1.21 (0, 2) |
| Communication (COMM, CCC) | 7.75 (4, 11) | 10.29 (5, 14) | 8.6 (4, 12) |
| Education (years) | 14.01 (12, 15) | 13.98 (12, 16) | 14 (12, 15) |
| **Maternal characteristics** |  |  |  |
| Age (yars) | 30.52 (27, 34) | 30.82 (28, 34) | 30.62 (28, 34) |
| ADHD (ADHD-RS) | 7.38 (5, 10) | 7.15 (5, 9) | 7.3 (5, 10) |
| Depression (SCL-5) | 2.53 (0, 4) | 2.43 (0, 3) | 2.5 (0, 3) |

ASD = Autism spectrum disorder, LD = Learning difficulties, Lang = Language problems, Beh = behavior problems, SEA = special educational assistance.
Abbreviations and original scales for PSD are given in parentheses (see methods section for full names). Numbers in parentheses are percent or first and third quartiles.

Inverse probability weights reduced the differences in mean values for covariates between participants followed up and those lost to follow up to less than 0.1 SMD (c.f Figure S1; [45]). Cumulative distribution plots showed that weighting balanced the entire distributions of covariates (Figures S7 and S8).

### Effects of special educational assistance

Consistent with the structural model shown in Figure 2, the analysis without adjustment showed that SEA at age five was associated with more psycho-social difficulties at age eight (c.f. Table S3 and Figure S7). Table S4 and Figures S9 and S10, S11, and S12 show coefficients of the adjusted regression model, which indicates that after adjustment for confounders SEA was associated with less psycho-social difficulties at age eight.

Over all psycho-social outcomes and groups of developmental or behavior problems the estimated average treatment effect (ATE) was a symptom reduction by 0.10 standardized mean deviations (SMD) (Credible Interval CI: 0.04, 0.16). Figure 3 shows that the 90% credible interval is for all groups above 0. The pairwise comparisons of all groups did not show clear differences in the estimated treatment effects between groups (c.f. Table S5 and Figure S14)

**Figure 3.**
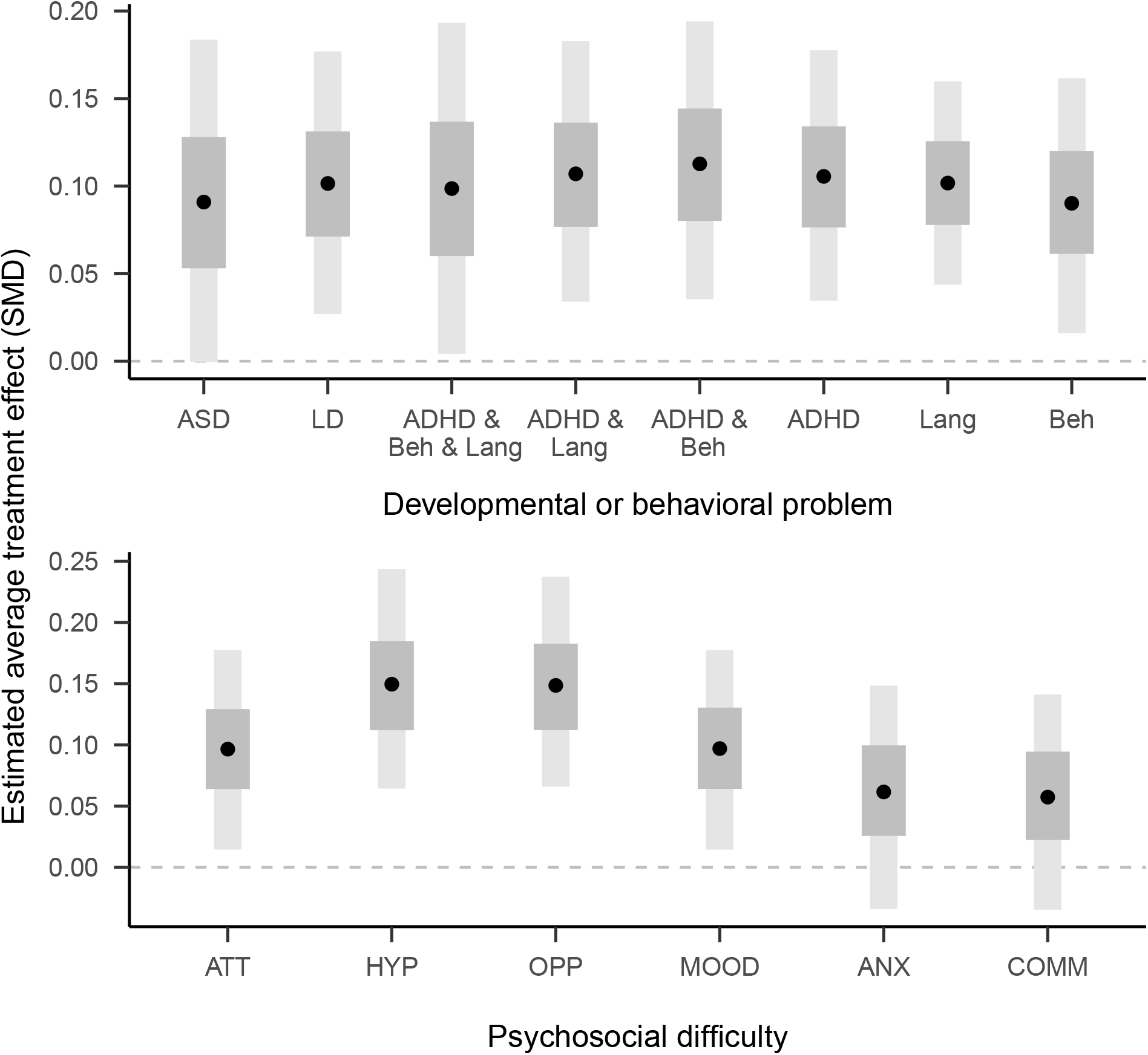
Estimated average treatment effects stratified by group (top) and outcome (bottom). Lines indicate means, gray and dark-gray bands indicate 50 and 95% credible intervals. SMD = standardized mean deviation. Abbreviations as in Table 1.

Figure 3 and Table 2 also show estimated effect sizes stratified by outcomes and indicate that SEA had a positive effect on all measured psycho-social outcomes. While there were some differences in the effect size estimates for different outcomes, in particular smaller effects for anxiety and communication problems, pairwise comparisons did not show reliable differences between them (c.f. Table S6 and Figure S15). Effect size estimates did not vary substantially by the child sex (c.f. Figure S18).

**Table 2.**
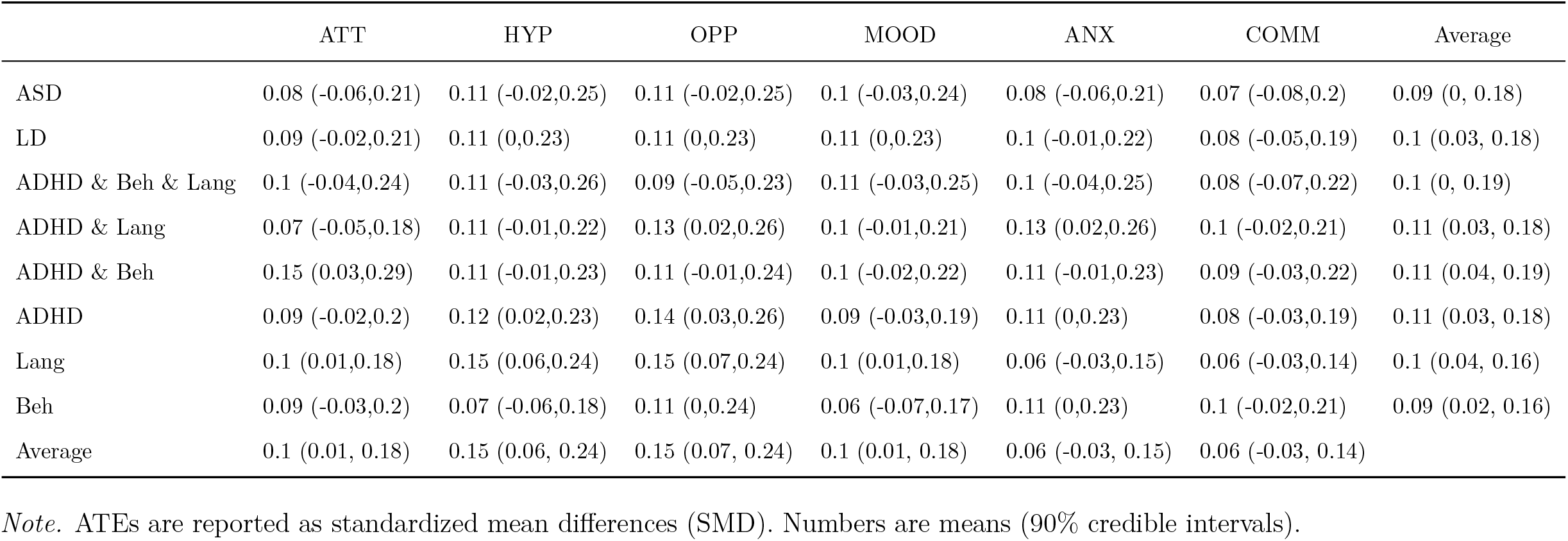
Estimated average treatment effects (ATE) stratified by groups with different developmental and behavioral problems (rows) and psycho-social difficulties (columns).

## Discussion

This research used observational data from a longitudinal population based cohort study to investigate the effect of special educational assistance (SEA) in ECEC on psycho-social difficulties of children with developmental or behavior problems. We found that, after adjustment for treatment indicators, mothers of children who received SEA in kindergarten reported fewer psycho-social difficulties three years later, compared to mothers whose children did not receive SEA.

While there was some variation in the extent of the positive effect of SEA between groups and different psycho-social difficulties, these differences were not reliably different from zero (c.f. Figures S14 and S15). Because the credible intervals for these differences are large compared to the magnitude of the estimated overall effect and the random effects standard deviations are clearly non-zero (S4), these results do not exclude the possibility of group differences. Instead, they might reflect difficulties in reliably measuring exposure, covariates, and outcomes based on parent reports only. Still, the available data were sufficient to reveal an overall positive effect of SEA.

While the positive effect reported in this study is consistent with the results of randomized controlled trials [10, 11] and with reports of the positive effects of preschool child care quality [46], it also stands in contrast to previous observational studies, which estimated no or a small negative “effects” of special education. This apparent contradiction can be due to a number of differences between the current and previous studies. We had estimates of pre-treatment difficulties, and could estimate effects of special education on the change of psycho-social difficulties. Moreover, we used adjustment for treatment predictors instead of propensity score weighting. Adjustment is the preferable approach if treatment-predictors are not colliders on a backdoor path from outcome to treatment and if the sample size is large enough to allow for inclusion of many of adjustment variables. Another important difference is that whereas previous studies focused on scholastic outcomes, we focused on the effect on psycho-social difficulties. This is a to date little examined but important outcome of SEA, because early psycho-social difficulties are associate with impaired functioning in adulthood [3]. Interestingly, the clear results of SEA on externalizing behavior suggests that in addition to helping children with DBP, it can also benefit their families by reducing disruptive behavior.

The estimated effect size for the reduction of psycho-social difficulties is with on average 0.10 standardized mean difference small. In comparison, previous meta analysis about school– or ECEC–based interventions found effect sizes of between -.3 and 1.3 SMD for children with or at risk for ADHD [47, 48] or SMD between 0.3 and 1.1 for children with autism [49]. Randomized trials of classroom management training for kindergarten teachers showed effect sizes similar to our results [Cohen’s d around 0.3 for high risk children at the nine-months follow up, 50]. It is possible that the small effect sizes we estimated are, in addition to above mentioned measurement problems, due to the fact the SEA was often provided by personnel with limited training, especially for children with typically less severe problems (c.f. Figure S5). More generally, the decentralized organization of the Educational and Psychological Counselling Service is likely to lead to a large variation in the implementation of SEA [51]. MoBa did not collect more detailed data about SEA, which could help to elucidate when it is most effective. Another possible explanation is that the composition of the study sample, which over-represents well-educated families compared to the population [37], leads to an underestimation of the true effect size, because well-educated parents could reduce children’s psycho-social difficulties even without SEA [52].

While the current study showed that mothers report fewer psycho-social difficulties in elementary school when their children received SEA in ECEC, a causal interpretation of this result as reflecting an effect of SEA rests on a number of assumptions encoded in Figure 2. One un-testable assumption is that there are no unmeasured confounders that predict both which children receive SEA and their developmental pathway. Even though the reported analysis includes obvious confounders, other unobserved confounder could still account for some of the positive association of SEA and psycho-social development. However, because RCTs of SEA and similar interventions typically report positive effects, and thus confirm a causal role of SEA, it appears unlikely that the effects estimated in this study are primarily due to confounding.

Given that the current study does not have more precise and detailed measures, future studies that assess outcomes through blinded raters or objective instruments and measure quality and quantity of the treatment more thoroughly are needed. Studies that better assess variations in treatment quality and content and use more representative samples will be useful to investigate reasons for the relatively small effects observed in the current study, and to identify criteria for effective interventions in ECEC.

## Conclusion

Previous RCTs about special educational assistance and teacher management programs showed that interventions in ECEC have a positive immediate impact for children with developmental or behavioral problems, but provide little guidance on long-term effects. The current study has due to its observational character a lower internal validity than RCTs, but complements them in terms of external validity and by examining long-term effects. It thus strengthens the view that interventions in ECEC are a useful approach to support preschoolers with developmental or behavioral problems.

In sum, the current study suggests that the psycho-social development of children with developmental or behavior problems can be modified in a positive way through interventions in ECEC, also when provided outside the structured context of randomized controlled trials. Future research with better measurements and more representative samples should investigate under which conditions such interventions are most effective.

## Key points and relevance

- Parent training programs are considered a key component of early interventions against the development of developmental or mental health problems
- Special educational assistances in early childhood education and care (SEA in ECEC) showed positive effects on later outcomes in RCTs, but population based cohort studies reported no or even negative associations
- Results from our large, population based cohort study indicate that SEA in ECEC is associated with reduced psycho-social difficulties in elementary school
- SEA in ECEC may be effective also when implemented outside the structured context of RCTs and for children who do not come from disadvantaged backgrounds
- Easy access to SEA in ECEC may be a component of early intervention strategies to prevent or mitigate development of psycho-social difficulties in preschoolers at risk.

## Supporting information

Supplementary methods and results

## Data Availability

Data are available upon application to the Norwegian Mother, Father and Child Cohort Study (MoBa)
https://www.fhi.no/en/studies/moba/

## References

1. Wichstrøm L, Berg-Nielsen TS, Angold A, et al (2012) Prevalence of psychiatric disorders in preschoolers. Journal of child psychology and psychiatry, and allied disciplines 53:695–705

2. Global Research on Developmental Disabilities Collaborators (2018) Developmental disabilities among children younger than 5 years in 195 countries and territories, 1990-2016: a systematic analysis for the Global Burden of Disease Study 2016. The Lancet Global health 6:e1100–e1121

3. Copeland WE, Wolke D, Shanahan L, Costello EJ (2015) Adult Functional Outcomes of Common Childhood Psychiatric Problems: A Prospective, Longitudinal Study. JAMA psychiatry 72:892–899

4. Knudsen EI, Heckman JJ, Cameron JL, Shonkoff JP (2006) Economic, neurobiological, and behavioral perspectives on building America’s future workforce. Proceedings of the National Academy of Sciences of the United States of America 103:10155–10162

5. Heckman JJ (2006) Skill formation and the economics of investing in disadvantaged children. Science 312:1900–1902

6. Reynolds AJ, Temple JA, Robertson DL, Mann EA (2001) Long-term effects of an early childhood intervention on educational achievement and juvenile arrest: A 15-year follow-up of low-income children in public schools. JAMA: the journal of the American Medical Association 285:2339–2346

7. Perrin EC, Sheldrick RC, McMenamy JM, et al (2014) Improving parenting skills for families of young children in pediatric settings: a randomized clinical trial. JAMA pediatrics 168:16–24

8. OECD (2019) Education at a Glance 2019: OECD Indicators. OECD Publishing, Paris

9. Hagen ÅM, Melby-Lervåg M, Lervåg A (2017) Improving language comprehension in preschool children with language difficulties: a cluster randomized trial. Journal of child psychology and psychiatry, and allied disciplines 58:1132–1140

10. Zwaigenbaum L, Bauman ML, Choueiri R, et al (2015) Early Intervention for Children With Autism Spectrum Disorder Under 3 Years of Age: Recommendations for Practice and Research. Pediatrics 136 Suppl 1:S60–81

11. Reinke WM, Herman KC, Dong N (2018) The Incredible Years Teacher Classroom Management Program: Outcomes from a Group Randomized Trial. Prevention science: the official journal of the Society for Prevention Research 19:1043–1054

12. Balzer LB (2017) “All Generalizations Are Dangerous, Even This One.”-Alexandre Dumas. Epidemiology 28:562–566

13. Cole SR, Stuart EA (2010) Generalizing evidence from randomized clinical trials to target populations: The ACTG 320 trial. American journal of epidemiology 172:107–115

14. Huitfeldt A, Stensrud MJ (2018) Re: Generalizing Study Results: A Potential Outcomes Perspective. Epidemiology 29:e13–e14

15. Lesko CR, Buchanan AL, Westreich D, et al (2017) Generalizing Study Results: A Potential Outcomes Perspective. Epidemiology 28:553–561

16. Sullivan AL, Field S (2013) Do preschool special education services make a difference in kindergarten reading and mathematics skills?: A propensity score weighting analysis. Journal of school psychology 51:243–260

17. Dempsey I, Valentine M, Colyvas K (2016) The Effects of Special Education Support on Young Australian School Students. International Journal of Disability, Development and Education 63:271–292

18. Wendelborg C, Caspersen J, Kittelsaa AM, et al (2015) Barnehagetilbudet til barn med særlige behov. NTNU Samfunnsforskning

19. Lekhal R (2018) Does special education predict students’ math and language skills? European journal of special needs education 33:525–540

20. Kvande MN, Bjørklund O, Lydersen S, et al (2018) Effects of special education on academic achievement and task motivation: a propensity-score and fixed-effects approach. European journal of special needs education 1–15

21. Morgan PL, Frisco M, Farkas G, Hibel J (2010) A Propensity Score Matching Analysis of the Effects of Special Education Services. The Journal of special education 43:236– 254

22. Magnus P, Birke C, Vejrup K, et al (2016) Cohort profile update: The norwegian mother and child cohort study (MoBa). International journal of epidemiology

23. Magnus P, Irgens LM, Haug K, et al (2006) Cohort profile: The norwegian mother and child cohort study (MoBa). International journal of epidemiology 35:1146–1150

24. Irgens LM (2000) The medical birth registry of norway. Epidemiological research and surveillance throughout 30 years. Acta obstetricia et gynecologica Scandinavica 79:435–439

25. Silva RR, Alpert M, Pouget E, et al (2005) A rating scale for disruptive behavior disorders, based on the DSM-IV item pool. The Psychiatric quarterly 76:327–339

26. Sharp C, Goodyer IM, Croudace TJ (2006) The Short Mood and Feelings Questionnaire (SMFQ): A Unidimensional Item Response Theory and Categorical Data Factor Analysis of Self-Report Ratings from a Community Sample of 7-through 11-Year-Old Children. Journal of abnormal child psychology 34:365–377

27. Birmaher B, Brent DA, Chiappetta L, et al (1999) Psychometric properties of the Screen for Child Anxiety Related Emotional Disorders (SCARED): a replication study. Journal of the American Academy of Child and Adolescent Psychiatry 38:1230–1236

28. Norbury CF, Nash M, Baird G, Bishop D (2004) Using a parental checklist to identify diagnostic groups in children with communication impairment: a validation of the Children’s Communication Checklist–2. International journal of language & communication disorders / Royal College of Speech & Language Therapists 39:345–364

29. Conners CK, Sitarenios G, Parker JDA, Epstein JN (1998) The Revised Conners’ Parent Rating Scale (CPRS-R): Factor Structure, Reliability, and Criterion Validity. Journal of abnormal child psychology 26:257–268

30. Nøvik TS (1999) Validity of the child behaviour checklist in a norwegian sample. European child & adolescent psychiatry 8:247–254

31. Achenbach TM, Ruffle TM (2000) The Child Behavior Checklist and related forms for assessing behavioral/emotional problems and competencies. Pediatrics in review / American Academy of Pediatrics 21:265–271

32. Kessler RC, Adler L, Ames M, et al (2005) The world health organization adult ADHD Self-Report scale (ASRS): A short screening scale for use in the general population. Psychological medicine 35:245–256

33. Strand BH, Dalgard OS, Tambs K, Rognerud M (2003) Measuring the mental health status of the norwegian population: A comparison of the instruments SCL-25, SCL-10, SCL-5 and MHI-5 (SF-36). Nordic journal of psychiatry 57:113–118

34. R Core Team (2017) R: A language and environment for statistical computing. R Foundation for Statistical Computing, Vienna, Austria

35. Bürkner P-C (2017) brms: An R package for Bayesian multilevel models using Stan. Journal of Statistical Software 80:1–28. https://doi.org/10.18637/jss.v080.i01

36. Greenland S, Pearl J, Robins JM (1999) Causal diagrams for epidemiologic research. Epidemiology 10:37–48

37. Biele G, Gustavson K, Czajkowski NO, et al (2019) Bias from self selection and loss to follow-up in prospective cohort studies. European journal of epidemiology 34:927–938

38. Stan Development Team (2018) RStan: The R interface to Stan

39. Carpenter B, Gelman A, Hoffman M, et al (2017) Stan: A probabilistic programming language. Journal of statistical software 76:1–32

40. Greenland S (2000) Principles of multilevel modelling. International journal of epidemiology 29:158–167

41. Gelman A, Hill J, Yajima M (2012) Why We (Usually) Don’t Have to Worry About Multiple Comparisons. Journal of research on educational effectiveness 5:189–211

42. Buuren S van, Groothuis-Oudshoorn K (2011) Mice: Multivariate imputation by chained equations in r. Journal of Statistical Software 45:1–67

43. Sullivan GM, Feinn R (2012) Using Effect Size-or Why the P Value Is Not Enough. Journal of graduate medical education 4:279–282

44. Wasserstein RL, Schirm AL, Lazar NA (2019) Moving to a World Beyond “p < 0.05”. The American statistician 73:1–19

45. Austin PC, Stuart EA (2015) Moving towards best practice when using inverse probability of treatment weighting (IPTW) using the propensity score to estimate causal treatment effects in observational studies. Statistics in medicine 34:3661–3679

46. Vandell DL, Belsky J, Burchinal M, et al (2010) Do effects of early child care extend to age 15 years? Results from the NICHD study of early child care and youth development. Child development 81:737–756

47. Richardson M, Moore DA, Gwernan-Jones R, et al (2015) Non-pharmacological interventions for attention-deficit/hyperactivity disorder (ADHD) delivered in school settings: Systematic reviews of quantitative and qualitative research. Health technology assessment 19:1–470

48. Gaastra GF, Groen Y, Tucha L, Tucha O (2016) The Effects of Classroom Interventions on Off-Task and Disruptive Classroom Behavior in Children with Symptoms of Attention-Deficit/Hyperactivity Disorder: A Meta-Analytic Review. PloS one 11:e0148841

49. Reichow B (2012) Overview of meta-analyses on early intensive behavioral intervention for young children with autism spectrum disorders. Journal of autism and developmental disorders 42:512–520

50. Fossum S, Handegård BH, Britt Drugli M (2017) The Incredible Years Teacher Classroom Management Programme in Kindergartens: Effects of a Universal Preventive Effort. Journal of child and family studies 26:2215–2223

51. Nilsen S, Herlofsen C (2012) National Regulations and Guidelines and the Local Follow-up in the Chain of Actions in Special Education. International journal of special education 27:136–147

52. Russell AE, Ford T, Russell G (2015) Socioeconomic Associations with ADHD: Findings from a Mediation Analysis. PloS one 10:e0128248

