## Supplementary methods and results for "The effect of special educational assistance in early childhood education and care on psycho-social difficulties in elementary school children"

### Supplementary Material

#### Supplementary Methods

All analysis were performed with R [Version 4.0.2; [1]][1]. Scripts for all analysis steps are available at https://github.com/gbiele/SPS358.

**Classification into groups with different developmental or behavioral problems (DBP).** Many children had combinations of developmental or behavioral problems. Altogether, we found 25 patterns of DBP, which we reduced to 8 non-overlapping groups. The guiding principle was to categorize each child according to it most severe problem, where the order of severity was ASD, LD, ADHD, language, and behavior problems. More specifically, we used following classification rules:

1. ASD = children with ASD (could have learning disabilities)
2. LD = children with learning disabilities, but no ASD
3. ADHDBehLang = children with ADHD and behavior and language problems but no ASD or LD,
4. ADHDLang = children with ADHD and language problems but no ASD, LD, or behavior problems,
5. ADHDBeh = children with ADHD and behavior problems but no ASD, LD, or

[1] We, furthermore, used the R-packages *apaTables* [Version 2.0.8; [2]], *arsenal* [Version 3.5.0; [3]], *bayesplot* [Version 1.8.0; [4]], *brms* [Version 2.14.4; [5]; [6]], *colorspace* [Version 2.0.0; [7]; [8]], *data.table* [Version 1.13.6; [9]], *DescTools* [Version 0.99.34; [10]], *english* [Version 1.2.5; [11]], *flextable* [Version 0.6.1; [12]], *Formula* [Version 1.2.3; [13]], *ggplot2* [Version 3.3.3; [14]], *ggpubr* [Version 0.4.0; [15]], *Gmisc* [Version 1.9.2; [16]], *Hmisc* [Version 4.4.0; [17]], *htmlTable* [Version 2.1.0; [18]], *jtools* [Version 2.1.0; [19]], *kableExtra* [Version 1.3.1; [20]], *knitr* [Version 1.30; [21]], *lattice* [Version 0.20.41; [22]], *magrittr* [Version 2.0.1; [23]], *officer* [Version 0.3.16; [24]], *papaja* [Version 0.1.0.9997; [25]], *pdftools* [Version 2.3.1; [**R-pdftools?**]], *Rcpp* [Version 1.0.6; [26]; [27]], *rstan* [Version 2.21.2; [28]; [29]], *rstanarm* [Version 2.21.1; [29]], *spelling* [30], *StanHeaders* [Version 2.21.0.6; [31]], and *survival* [Version 3.2.7; [32]].

language problems,

6. ADHD = children with ADHD problems but no ASD, LD, behavior or language problems,
7. Lang = children with language problems but no ASD, LD, ADHD or language problems,
8. Beh = children with oppositional defiant and/or conduct disorder problems (but no ASD, LD, ADHD or language problems)

Table S1 shows to which categories children with different developmental difficulties were assigned.

Table S1

*Patterns of developmental and behavioral problems and their classification into groups*

| | ASD | LD | ALB | AL | AB | ADHD | Lang | Beh |
|---|---|---|---|---|---|---|---|---|
| ADHD | 0 | 0 | 0 | 0 | 0 | 401 | 0 | 0 |
| ADHD + ASD | 2 | 0 | 0 | 0 | 0 | 0 | 0 | 0 |
| ADHD + Beh | 0 | 0 | 0 | 0 | 146 | 0 | 0 | 0 |
| ADHD + Beh + ASD | 1 | 0 | 0 | 0 | 0 | 0 | 0 | 0 |
| ADHD + Beh + ASD + LD | 1 | 0 | 0 | 0 | 0 | 0 | 0 | 0 |
| ADHD + Beh + Lang | 0 | 0 | 31 | 0 | 0 | 0 | 0 | 0 |
| ADHD + Beh + Lang + ASD | 3 | 0 | 0 | 0 | 0 | 0 | 0 | 0 |
| ADHD + Beh + Lang + ASD + LD | 2 | 0 | 0 | 0 | 0 | 0 | 0 | 0 |
| ADHD + Beh + Lang + LD | 0 | 9 | 0 | 0 | 0 | 0 | 0 | 0 |
| ADHD + Beh + LD | 0 | 2 | 0 | 0 | 0 | 0 | 0 | 0 |
| ADHD + Lang | 0 | 0 | 0 | 143 | 0 | 0 | 0 | 0 |
| ADHD + Lang + ASD | 8 | 0 | 0 | 0 | 0 | 0 | 0 | 0 |
| ADHD + Lang + ASD + LD | 3 | 0 | 0 | 0 | 0 | 0 | 0 | 0 |
| ADHD + Lang + LD | 0 | 24 | 0 | 0 | 0 | 0 | 0 | 0 |
| ADHD + LD | 0 | 4 | 0 | 0 | 0 | 0 | 0 | 0 |
| ASD | 8 | 0 | 0 | 0 | 0 | 0 | 0 | 0 |
| ASD + LD | 1 | 0 | 0 | 0 | 0 | 0 | 0 | 0 |
| Beh | 0 | 0 | 0 | 0 | 0 | 0 | 0 | 320 |
| Beh + ASD | 5 | 0 | 0 | 0 | 0 | 0 | 0 | 0 |
| Beh + Lang | 0 | 0 | 0 | 0 | 0 | 0 | 26 | 0 |
| Lang | 0 | 0 | 0 | 0 | 0 | 0 | 1307 | 0 |

Table S1 continued

| | ASD | LD | ALB | AL | AB | ADHD | Lang | Beh |
|---|---|---|---|---|---|---|---|---|
| Lang + ASD | 7 | 0 | 0 | 0 | 0 | 0 | 0 | 0 |
| Lang + ASD + LD | 2 | 0 | 0 | 0 | 0 | 0 | 0 | 0 |
| Lang + LD | 0 | 28 | 0 | 0 | 0 | 0 | 0 | 0 |
| LD | 0 | 15 | 0 | 0 | 0 | 0 | 0 | 0 |

*Note.* Cells show number of cases. ALB = ADHD & Lang & Beh, AL = ADHD & Lang, AB = ADHD & Beh. All other appreviations as in Table 1 of the main article

**Multiple chained imputation of missing values.** Table S2 shows the percentage of missing data. The analysis used altogether 214 variables, of which 172 were items assessing psycho-social difficulties.

To deal with missing data among co-variates, we generated 50 data sets with imputed missing data using the multiple chained imputation approach as implemented in the R package mice [33]. The imputation data set included 3662 participants for which the five or eight year questionnaire was available. The rating data was imputed on the level of individual rating items, which lead to a large number of 214 variables for the imputation process. Because the data set included 187 ordinal variables (mostly rating scale items) and the mice package does currently not offer facilities for fast imputation of ordinal data, we wrote a mice extension (https://github.com/gbiele/spolr) for efficient calculation of penalized ordered, logistic, and Gaussian regression using the rstan package. Here, penalization is implemented by putting weakly informative priors on regression weights for z-standardized predictors and estimating maximum a-posteriori parameter estimates.

We ran mice for four chains with 100 iterations each and verified through visual inspection that all chains had converged.

**Inverse probability of continued participation weights.** To predict loss to follow up from the MoBa age five to eight year questionnaire we used following groups of variables :

- child psycho-social difficulties at age 5
- maternal mental health
- type of the developmental or behavioral problem
- maternal age and education
- parity, birth month, child gender
- contact with health services
- special educational assistance (see also Figure S1)

Consistent with recommendations for calculating weights for hierarchical analysis, we used a hierarchical regression analysis [implemented in rstanarm, 34] as the basis for calculating stabilized inverse probability of continued participation weights (IPPW) for groups with different DBPs. In particular, we estimated a hierarchical logistic regression model with random intercepts and slopes, with type of developmental or behavioral problem, and child gender as grouping variables for random/group-specific effects. We fit a selection model for each of the 50 imputed data in R, using rstanarm [35], and obtained inverse probability of continued participation weights as the continuation probability for all participants in the 5 year questionnaire divided by each individuals continuation probability predicted by the selection model.

When evaluating inverse probability of continued participation weights, the focus should be on the successful balancing of the weighted follow up sample with the original inclusion sample, with respect to potential confounders [36]. Figure S1 shows standardized mean deviations (SMD) for all imputed samples, once non-weighted and once weighted by inverse probability of participation weights. A conventional threshold is that the magnitude of the SMD between participants who were and were not lost to follow up should be below

0.1. In the Figure, dots to the right (left) of the vertical zero line indicate that participants with high values on the variable were more (less) likely to also participate in the 8 year questionnaire.

In addition to means, the entire distributions of potential confounders should be matched. This is typically investigate through visual inspection of cumulative density plots, which are shown in Figure S2. Figure S3 directly shows the difference of the empirical cumulative distribution function and reinforces that weighting reduces the differences in the distribution of values for all predictor variables.

Table S2

*Percent missing data*

| | Participants with MoBa age five, eight data | | | Participants with at least MoBa age eight data | | |
|---|---|---|---|---|---|---|
| | all variables | age 5 scales | age 8 scales | all variables | age 5 scales | age 8 scales |
| Minimum | 0.0 | 0.2 | 0.3 | 0.0 | 0.5 | 31.9 |
| 1st Quartile | 0.6 | 0.7 | 0.5 | 0.9 | 0.7 | 32.1 |
| Median | 0.8 | 0.8 | 0.6 | 8.9 | 0.9 | 32.2 |
| Mean | 3.1 | 1.9 | 0.7 | 15.0 | 2.0 | 32.2 |
| 3rd. Quartile | 1.8 | 1.0 | 0.8 | 32.1 | 1.3 | 32.3 |
| Maximum | 65.7 | 65.7 | 1.4 | 67.2 | 67.2 | 32.7 |

*Note.* Age 5/8 scales refers to items to assess psycho-social difficulties. MoBa = Norwegian Mother, Father and Child Cohort Study

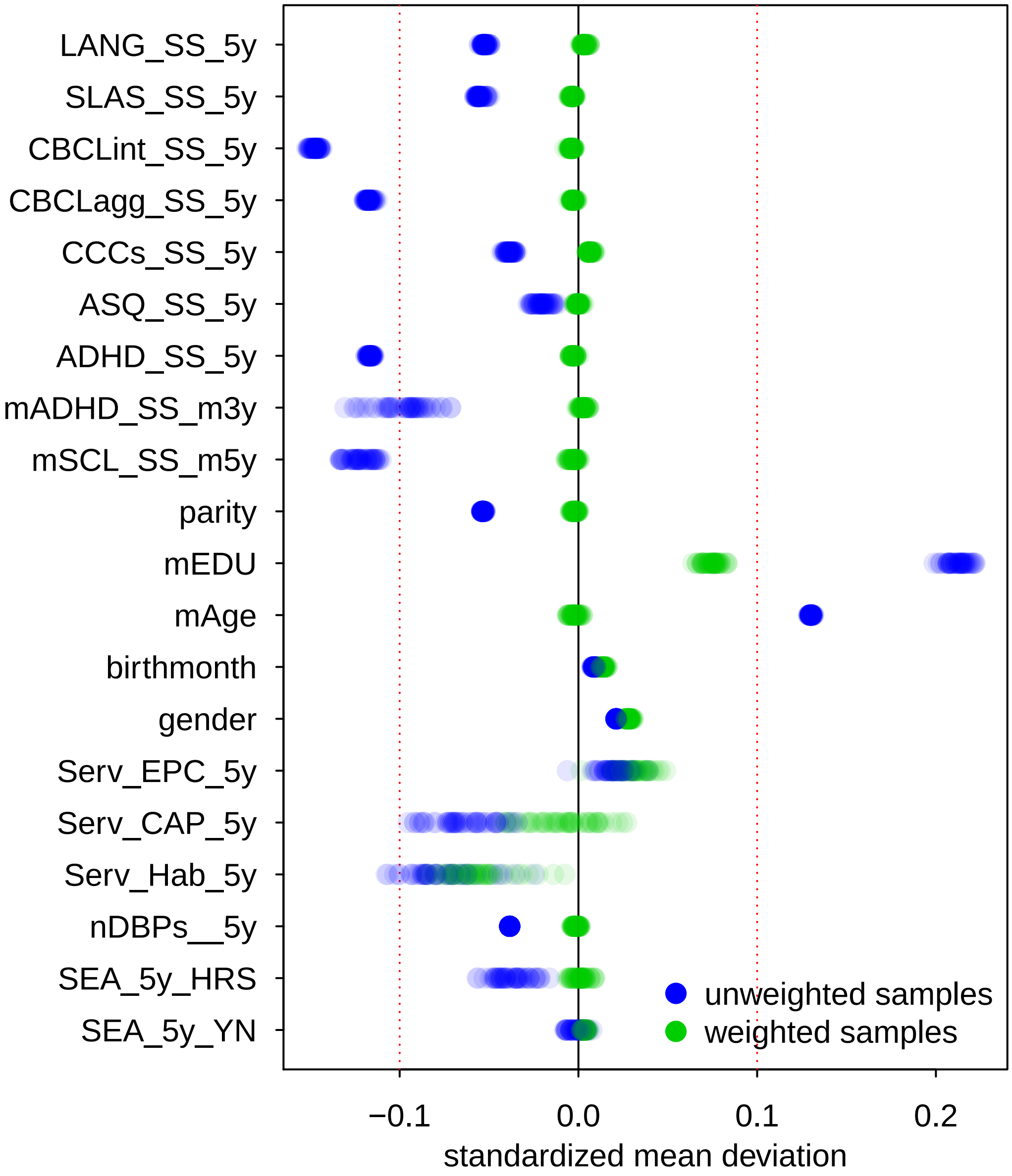


*Figure S1*. Standardized mean difference between respondents to the age 8 year questionnaire and those lost to follow up. For each variable that influences participation, each sample is represented with 50 transparent, overlapping dots (one for each imputed data set) for the standardized mean difference (SMD) between participants who were and were not lost to follow up. SS = sum-score, 5y = 5 year questionnaire, nDBP = number of developmental of behavioral problems, EPC = educational and psychological counseling service, CAP = Child and adolescent psychiatric units.

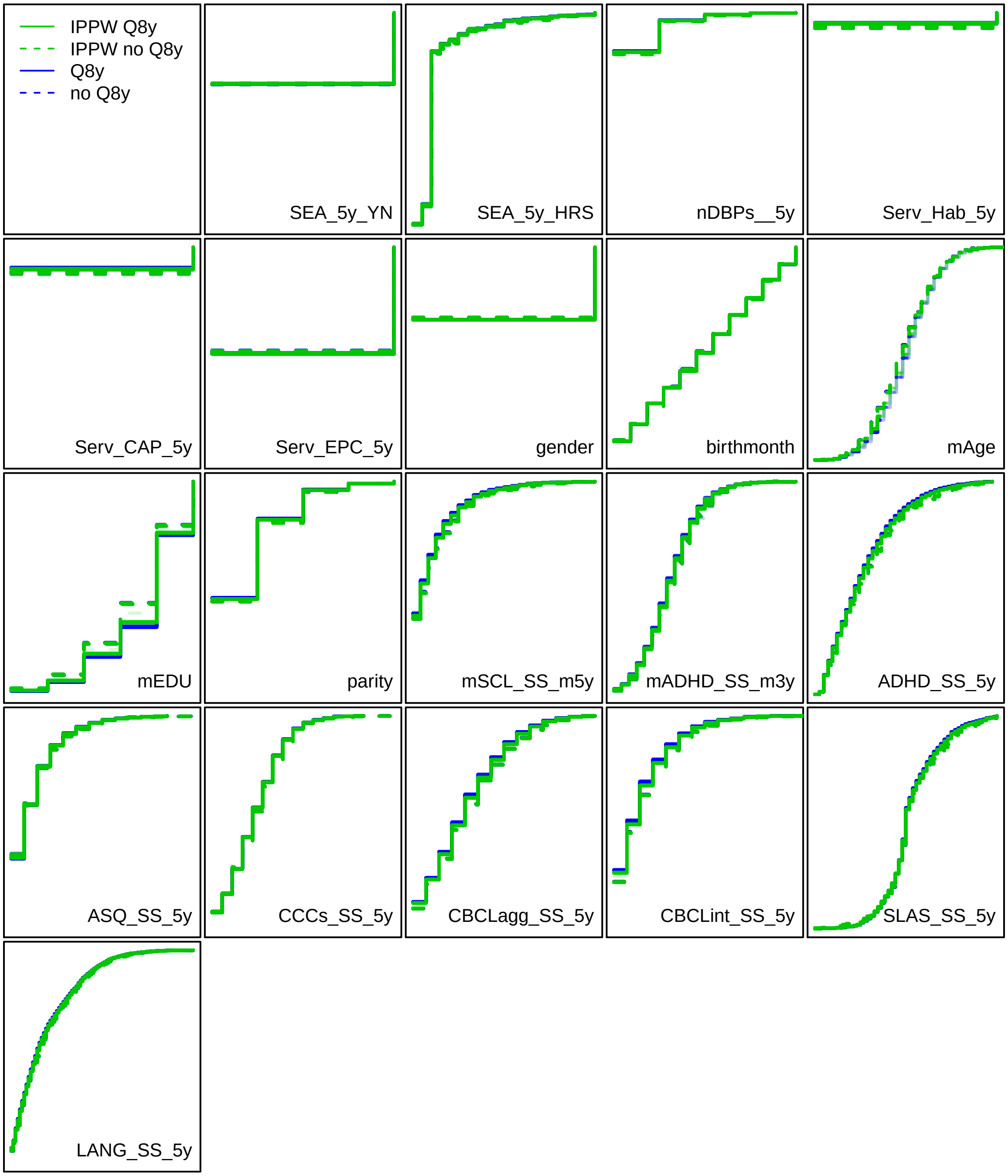


*Figure S2.* Cumulative distributions for respondents to the 8 year questionnaire (Q8y) and those lost to follow up (no Q8y) before weighting (blue) and after weighting (green). Increasing values of participation predictors are on the x-axis, the cumulative proportion of participants up to a variable value are on the y-axis. The sample is properly balanced, if the cumulative distribution function for participants with and without 8 year questionnaire are identical (i.e. the slid and dashed lines overlap).

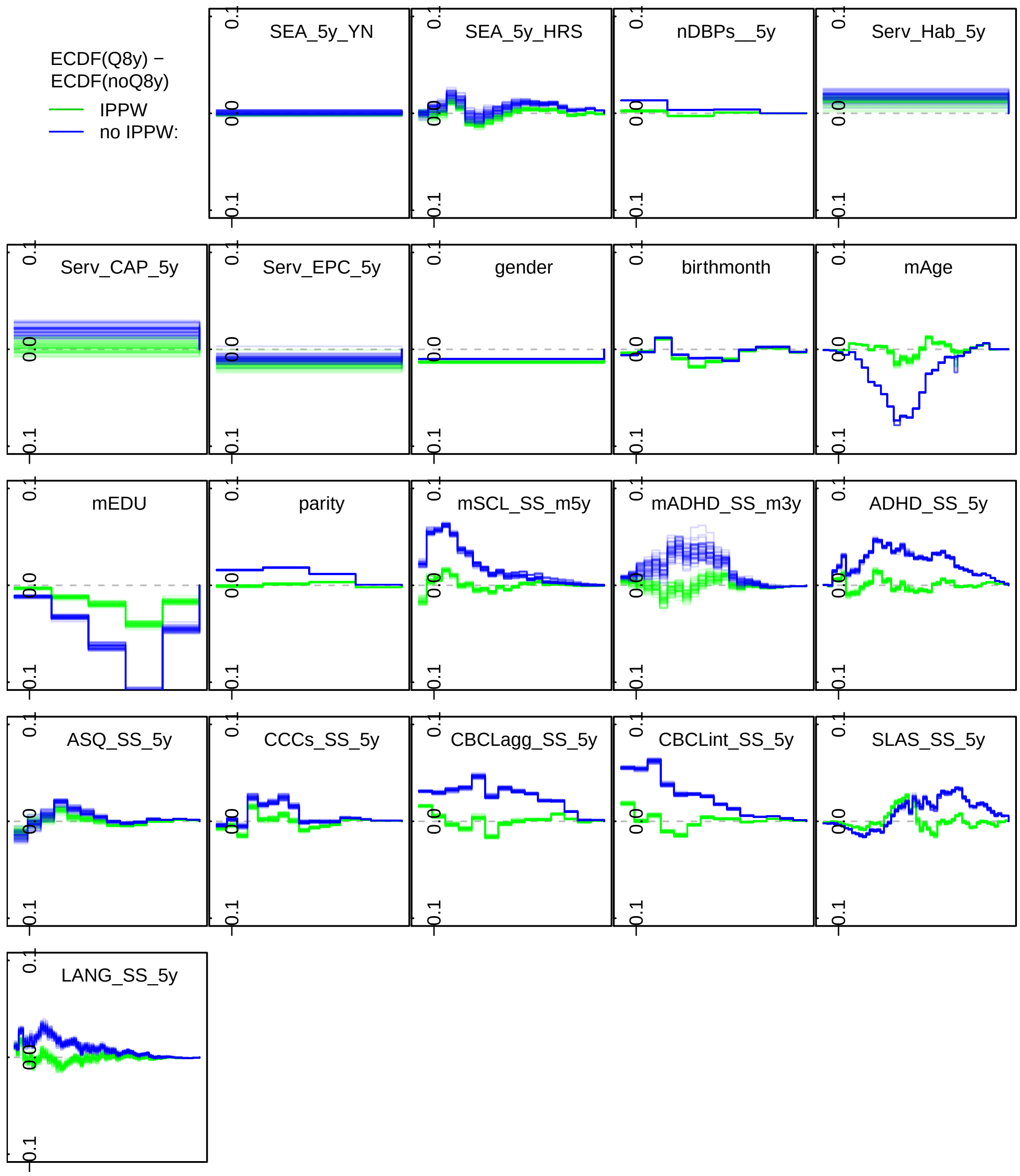


*Figure S3*. Difference in cumulative distributions for respondents to the 8 year questionnaire (Q8y) and those lost to follow up (no Q8y) before weighting (blue) and after weighting (green). Increasing values of participation predictors are on the Z axis, the differences of the cumulative proportion of participants up to a variable value are on the y axis. The sample is properly balanced, if the differences between participants with and without 8 year questionnaire is close to zero.

**Regression models and estimation of average treatment effects.** Outcome variables are sum-scores, which we model as ordinal variables. That is, the analysis model assumes latent traits for psycho-social difficulties, which in combination with cut K-1 cut points result in one of K possible sum-scores. This approach captures the intuition that sum-scores from questionnaires do not measure psycho-social difficulties on an interval scale and it also facilitates calculation of average treatment effects on the scale of standardized mean differences.

In order to obtain reliable estimates also for smaller groups, and in order to deal with the multiple comparison problem, we estimated the effects for all groups and psycho-social difficulties jointly in a hierarchical regression.

In particular, we estimate a random-intercept random-slope model for strata determined by (a) developmental or behavioral problems, (b) psycho-social difficulties, and (c) sex Further, we account for repeated measures within individuals by estimating individual level intercepts/random effects. We estimated the parameters for this model with a version of the (cumulative) ordinal regression implemented in the brms package[37, 38], which we modified to account for the fact that different outcome measures had different numbers of categories.

***Un-adjusted model.***

Even though the unadjusted model cannot be used to obtained unbiased effect estimate, we estimated such a model for reasons of completeness and to conform with STROBE reporting guidelines.

The unadjusted model estimated the effect of SEA as

$$
\begin{aligned}
sumscore \sim & 1 + SEA + h_{SEA} + h^2_{SEA} + \\
& (1 + SEA + h_{SEA} + h^2_{SEA} | DBP : PSD : sex) + \\
& (1|ID)
\end{aligned}
$$

where $SEA + h_{SEA} + h^2_{SEA}$ are the fixed effects for SEA and the linear and quadratic effects of the number of hours SEA, $(SEA + h_{SEA} + h^2_{SEA}|DBP : PSD : sex)$ are random effects for strata define by the type of developmental or behavior problem, outcome, and gender. $(1|ID)$ are participant level random effects.

The estimation of the model used inverse probability of continued participation weights to account for loss to follow up.

All non-binary predictors (including linear and quadratic effects) were scaled to mean of zero and a standard deviation of 1. We employed weakly informative shrinkage priors (normal distribution with a mean of zero and a standard deviation of 2) for the estimation of the fixed effects and for the estimation of the random effects standard deviation (half-normal distribution with a mean of zero and a standard deviation of 3). To verify convergence of all models we checked that all $\hat{R}$ values were below 1.1[39], and that the model estimation was completed without divergent iterations.

For each of the 50 imputed data sets, we estimated one model with four chains, each with 1000 warmup iteration and 500 post-warmup iterations. The reported results are based on the pooled results over all these analyses, i.e. based on 500 * 4 * 50 = 100,000 post warmup samples.

***Adjusted model.***

The basic structure of the adjusted model is the same as described for the un-adjusted model, except that we used additional covariates.

Figure 2 shows the causal assumptions underlying our analysis. Based on these causal assumptions we control bias due to treatment by indication by adjusting for maternal education and the degree of psycho-social difficulties at age five. Regarding the latter, we adjust for linear and quadratic effects of sum-scores for ADHD symptoms, externalizing and internalizing behavior (CBCL) and communication (CCC), developmental difficulties (ASQ) as well as interactions between these variables. We also included the number of developmental or behavioral problems at age five and contact to different types of mental health services as indicators of psycho-social difficulties at age five. We further adjusted for pregnancy duration, maternal education and mental health (ADHD and depression symptoms) as potential confounders, as well as for birth month and birth order, which are not of interest for the current study, but were shown to be associated with child mental health problems [40]. Lastly, we also adjusted for the number of children in the kindergarten group and the number of children per adult as indicators for variations in the quality of ECEC care.

***Calculation of average treatment effects.***

We calculated average treatment effects as

$$
\begin{aligned}
ATE_{g,o} =& \frac{\sum_{i=1}^{n_g} \omega_i TE_i}{\sum_{i=1}^{n_g} \omega_i} \\
TE =& \hat{Y}_1 - \hat{Y}_0 \\
\hat{Y}_1 =& \beta_F X_1 + \beta_R z_1 \\
\hat{Y}_0 =& \beta_F X_0 + \beta_R z_0
\end{aligned}
$$

where $ATE_{g,o}$ is the average treatment effect for a particular group and outcome, $\omega$ is a vector or IPPW weights, and $TE$ is a vector of treatment effects and $n_g$ is the number of individuals in a group. $X$ is a matrix with fixed effects predictors and $\beta_F$ are the associated regression weights. $z$ are random effects predictors and $\beta_R$ are the associated regression weights. In $X_1$ and $z_1$ the value for SEA is set to 1, and the value for the hours SEA is set to the true value for children who received SEA and to the imputed values for children who did

Table S3

*Coefficients of the hierarchical regression model without adjustment*

| predictor | FE coefficient | RE stand. dev. |
|---|---|---|
| supp_5y_YN | 1.053 (0.607, 1.513) | 2.111 (1.829, 2.456) |
| supp_5y_HRS.L | -0.142 (-0.287, 0) | 0.592 (0.503, 0.696) |
| supp_5y_HRS.Q | 0.174 (0.055, 0.297) | 0.52 (0.437, 0.619) |

*Note.* supp_5y_YN = Special educational assistance (SEA) received, supp_5y_HRS = hours SEA per week, FE = fixed effects, RE = random effects. L = linear effects, Q = quadratic effects. Numbers are means and 90% credible intervals.

not receive SEA. In $X_0$ and $z_0$ the values for SEA and hours SEA is set to 0. For reasons of transparency this description omits indices for the 50 imputations and 500 post warm-up MCMC samples per imputations, but all ATEs were calculated by averaging over all imputations and MCMC samples.

#### Supplementary Results

**Unadjusted model: The association between SEA and psycho-social and developmental problems.** The unadjusted model estimated a hierarchical regression model with only the presence of SEA and linear and quadratic effects of the number of hours SEA per week as predictors. Given the causal relationships displayed in Figure 2 the positive association between SEA and psycho-social and developmental problems at age 8 was expected. This should however not be taken as evidence a negative effect of SEA.

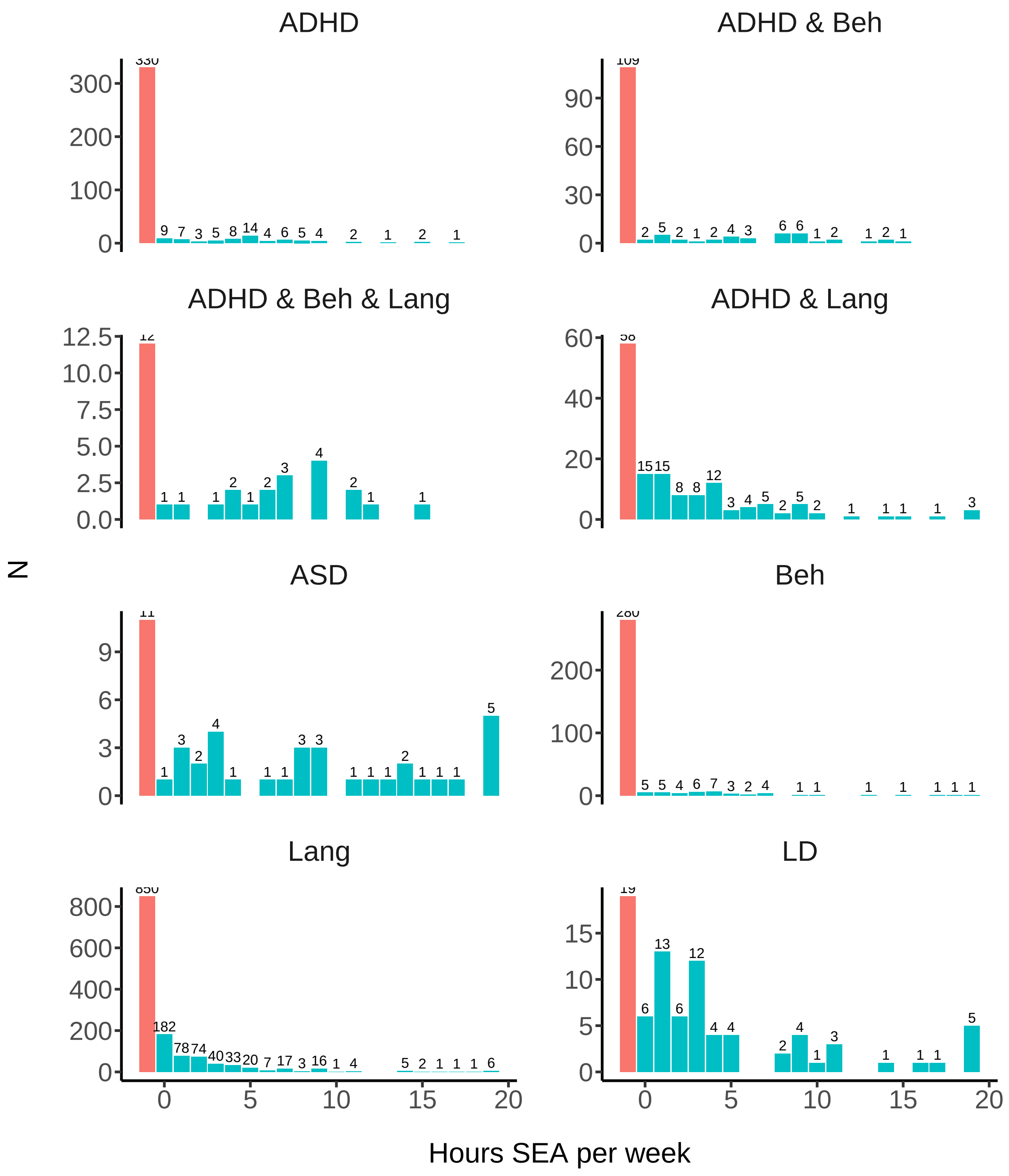


*Figure S4*. Distribution of weakly hours special educational assistance (SEA), stratified by type of developmental or behavioral problem.

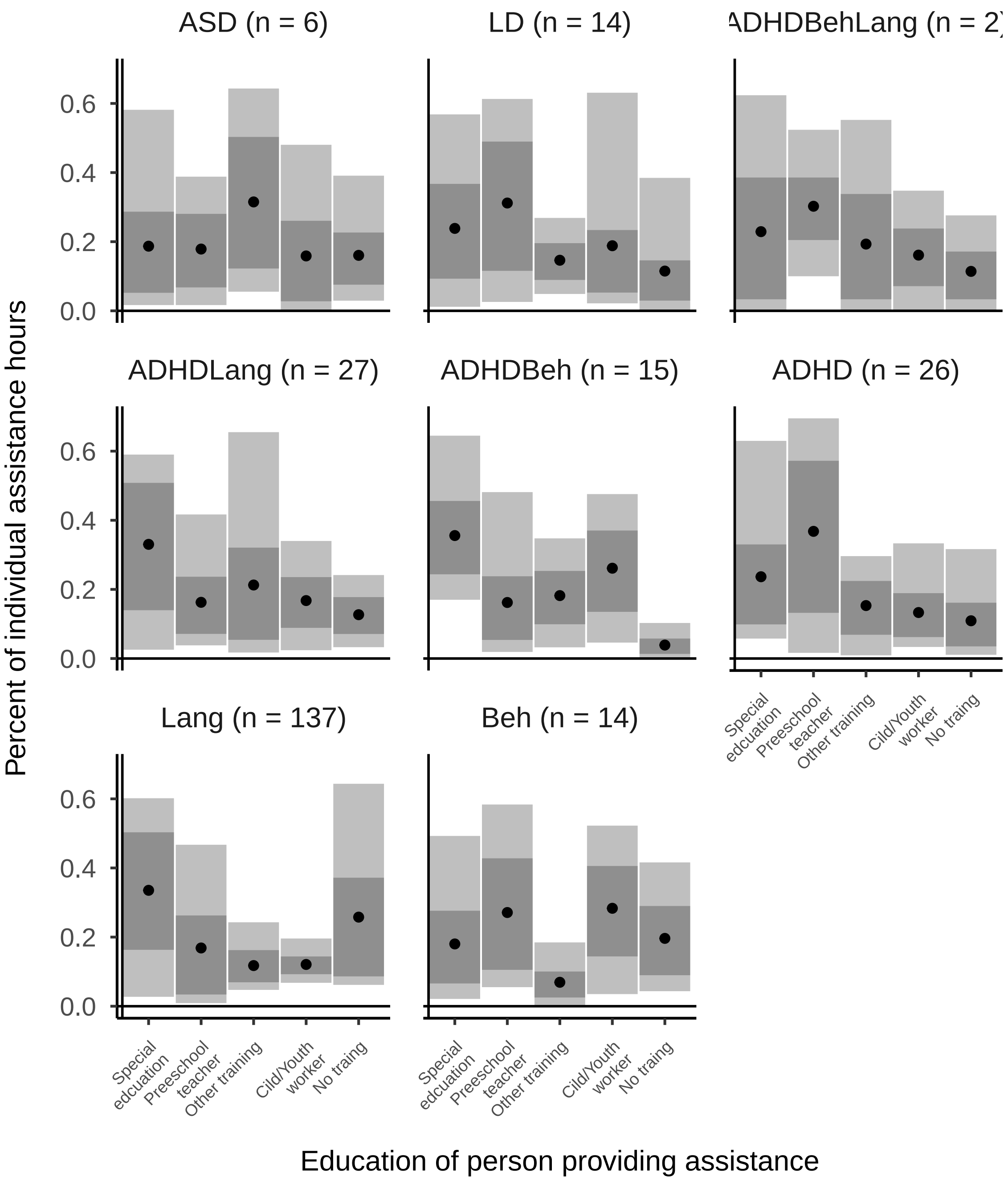


*Figure S5*. Education of employees providing individual assistance in ECEC to children with DBP. Only employees in the categories “Special education” and “Preschool teacher” have a longer pedagogical education. Only children with ASD, delayed development or combined ADHD and language problems receive the majority of assistance from persons with training in special education. All other children receive assistance mostly from regular preschool teachers or adults without dedicated education. Data are from the MoBa Kindergarten questionnaire, which was sent to and returned from only a subsample of the MoBa population.

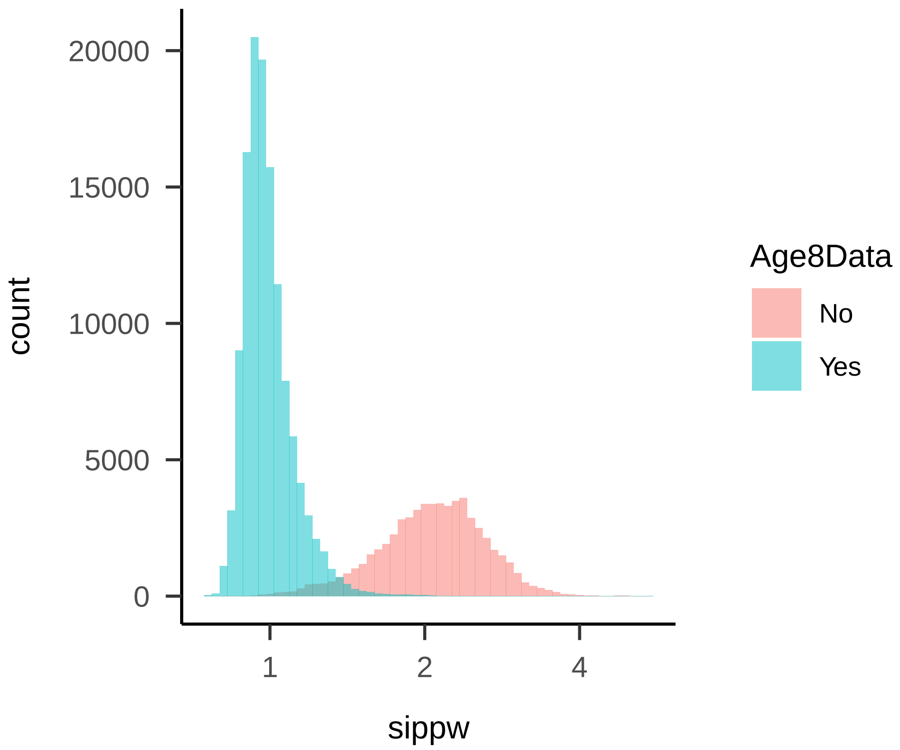


*Figure S6*. Distribution of stabilized inverse probability of continued participation weights (IPPW). The histogram shows weights for all 50 imputed data sets. Weights for participants with data from the age eight questionnaire range between 0.76 and 2.27.

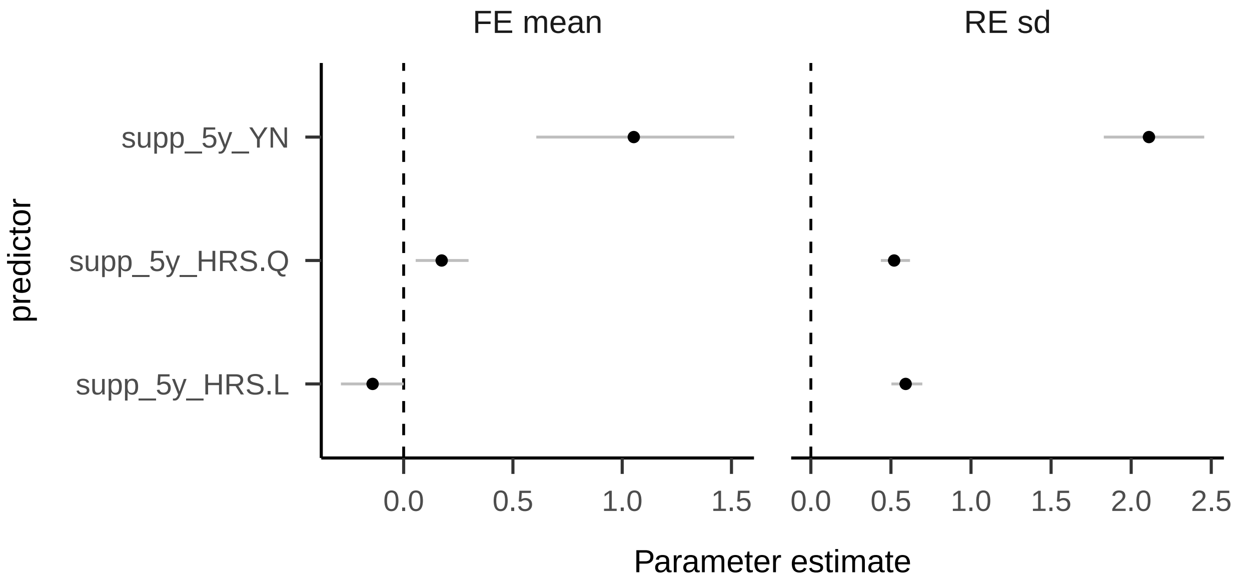


*Figure S7.* Coefficients of the hierarchical regression model without adjustment. FE mean = fixed effects, RE sd = standard deviation of random effects. L = linear effects, Q = quadratic effects. Colons between variables indicate interactions. The plot shows means as dots and 90% credible intervals as vertical lines.

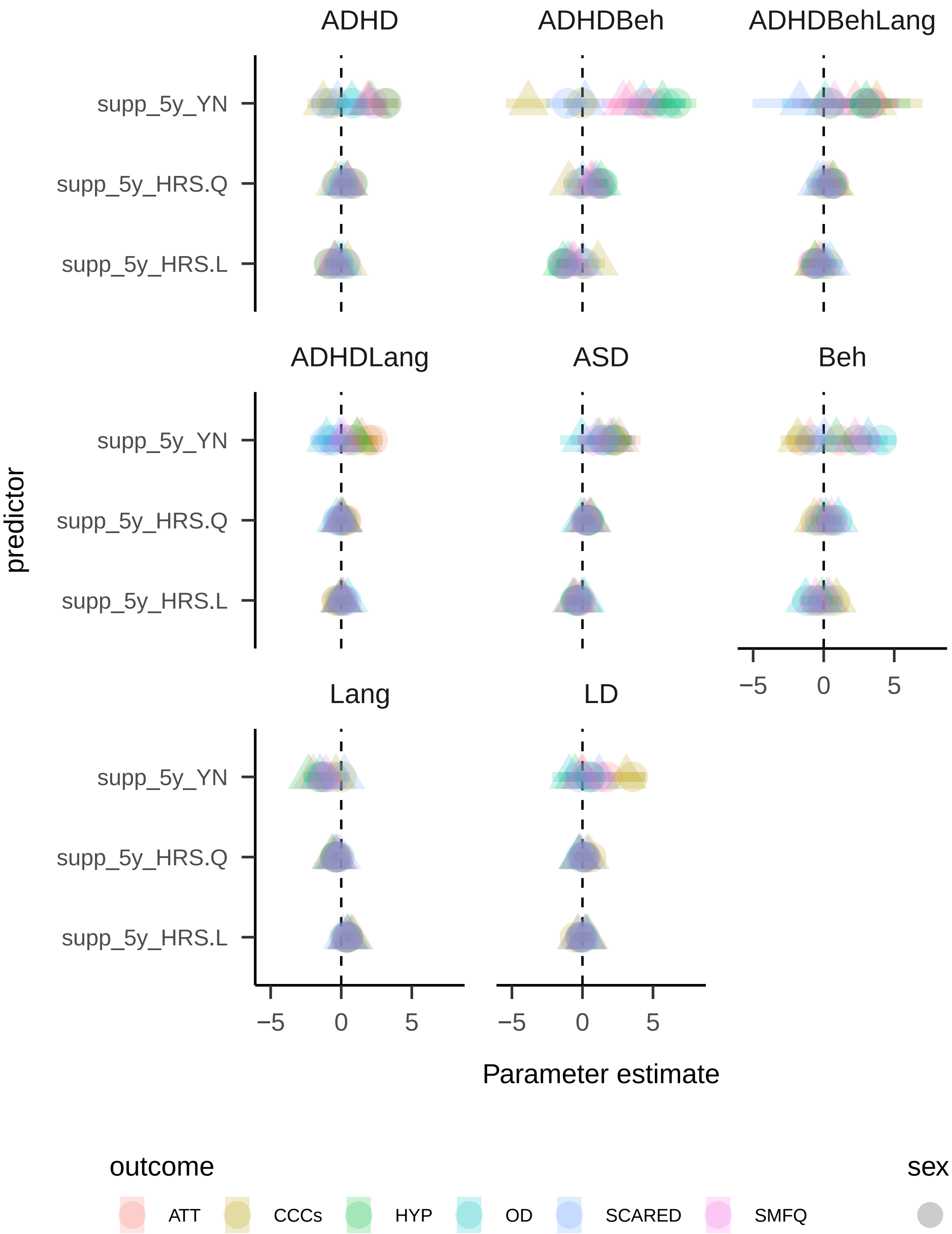


*Figure S8*. Group specific regression coefficients for the model without adjustment (fixed effects + random effects)

Table S4

*Coefficients of the hierarchical regression model with adjustment*

| predictor | FE coefficient | RE stand. dev. |
|---|---|---|
| supp_5y_YN | -0.213 (-0.391, -0.037) | 0.126 (0.018, 0.261) |
| supp_5y_HRS.L | 0.014 (-0.063, 0.09) | 0.036 (0.003, 0.093) |
| supp_5y_HRS.Q | 0.005 (-0.056, 0.067) | 0.027 (0.003, 0.076) |
| mEDU.L | -0.056 (-0.108, -0.004) | 0.1 (0.06, 0.145) |
| mEDU.Q | 0.02 (-0.027, 0.067) | 0.057 (0.01, 0.105) |
| nMHPs_5y.L | 0.055 (-0.029, 0.138) | 0.086 (0.009, 0.193) |
| nMHPs_5y.Q | -0.081 (-0.145, -0.018) | 0.059 (0.007, 0.131) |
| parity.L | -0.095 (-0.152, -0.037) | 0.138 (0.086, 0.196) |
| parity.Q | 0.056 (0.01, 0.102) | 0.024 (0.002, 0.069) |
| preg_week.L | 0.017 (-0.028, 0.063) | 0.03 (0.003, 0.079) |
| preg_week.Q | -0.007 (-0.051, 0.038) | 0.025 (0.002, 0.068) |
| birthmonth.L | 0.05 (-0.003, 0.102) | 0.113 (0.07, 0.162) |
| birthmonth.Q | -0.002 (-0.048, 0.044) | 0.055 (0.009, 0.107) |
| LNX_SS_5y.L | 0.371 (0.243, 0.499) | 0.573 (0.496, 0.665) |
| LNX_SS_5y.Q | -0.033 (-0.096, 0.031) | 0.099 (0.028, 0.166) |
| CBCLagg_SS_5y.L | 0.437 (0.337, 0.539) | 0.353 (0.292, 0.425) |
| CBCLagg_SS_5y.Q | 0.029 (-0.037, 0.095) | 0.036 (0.003, 0.096) |
| CBCLint_SS_5y.L | 0.248 (0.168, 0.329) | 0.248 (0.202, 0.302) |
| CBCLint_SS_5y.Q | -0.016 (-0.073, 0.041) | 0.102 (0.053, 0.149) |
| ATT_SS_5y.L | 0.24 (0.112, 0.368) | 0.388 (0.307, 0.479) |
| ATT_SS_5y.Q | -0.058 (-0.163, 0.048) | 0.144 (0.042, 0.226) |
| HYP_SS_5y.L | 0.261 (0.144, 0.378) | 0.273 (0.209, 0.349) |
| HYP_SS_5y.Q | 0.042 (-0.057, 0.142) | 0.106 (0.023, 0.178) |
| mADHD_SS_m3y.L | 0.109 (0.052, 0.166) | 0.102 (0.045, 0.159) |
| mADHD_SS_m3y.Q | -0.066 (-0.115, -0.016) | 0.063 (0.013, 0.116) |
| mSCL_SS_m5y.L | 0.094 (0.042, 0.147) | 0.068 (0.01, 0.129) |
| mSCL_SS_m5y.Q | -0.027 (-0.079, 0.024) | 0.081 (0.011, 0.163) |
| Serv_BUP_5yJa | 0.098 (-0.093, 0.293) | 0.185 (0.02, 0.41) |
| Serv_Hab_5yJa | 0.177 (-0.106, 0.452) | 0.217 (0.022, 0.542) |

Table S4 continued

| predictor | FE coefficient | RE stand. dev. |
|---|---|---|
| Serv_PPT_5yJa | 0.034 (-0.073, 0.14) | 0.08 (0.008, 0.199) |
| lnum_childr_KgGr | -0.132 (-0.291, 0.026) | 0.123 (0.064, 0.171) |
| num_childr_per_adult_KgGr | 0.037 (-0.006, 0.081) | 0.018 (0.002, 0.05) |
| LNX_SS_5y.L:CBCLagg_SS_5y.L | -0.051 (-0.116, 0.015) | 0.105 (0.035, 0.164) |
| LNX_SS_5y.L:CBCLint_SS_5y.L | -0.024 (-0.075, 0.027) | 0.024 (0.002, 0.066) |
| LNX_SS_5y.L:ATT_SS_5y.L | 0.029 (-0.052, 0.109) | 0.034 (0.003, 0.094) |
| LNX_SS_5y.L:HYP_SS_5y.L | 0.007 (-0.069, 0.083) | 0.061 (0.007, 0.131) |
| CBCLagg_SS_5y.L:CBCLint_SS_5y.L | -0.079 (-0.141, -0.018) | 0.058 (0.008, 0.111) |
| CBCLagg_SS_5y.L:ATT_SS_5y.L | -0.022 (-0.112, 0.066) | 0.034 (0.003, 0.096) |
| CBCLagg_SS_5y.L:HYP_SS_5y.L | -0.071 (-0.157, 0.016) | 0.028 (0.003, 0.082) |
| CBCLint_SS_5y.L:ATT_SS_5y.L | 0.056 (-0.02, 0.131) | 0.039 (0.004, 0.1) |
| CBCLint_SS_5y.L:HYP_SS_5y.L | 0.052 (-0.02, 0.125) | 0.028 (0.002, 0.079) |
| ATT_SS_5y.L:HYP_SS_5y.L | -0.046 (-0.162, 0.07) | 0.077 (0.009, 0.154) |

*Note.* supp_5y_YN = Special educational assistance (SEA) received, supp_5y_HRS = hours SEA per week, nMHPs = number of developmental or behavior problems, LNX = composite sum score for language difficulties, mADHD / mSCL = maternal ADHD and depressive symptoms, Serv_BUP/PPT/Hab = contact with child and adolescent psychiatric unit/educational and psychological counselling service, habilitation service, num_childr_KgGr = number of children in kindergten group, FE = fixed effects, RE = random effects. L = linear effects, Q = quadratic effects. 3y/5y = measured with MoBa age three/five years questionnaires. Colons between variables indicate interactions. Numbers are means and 90% credible intervals.

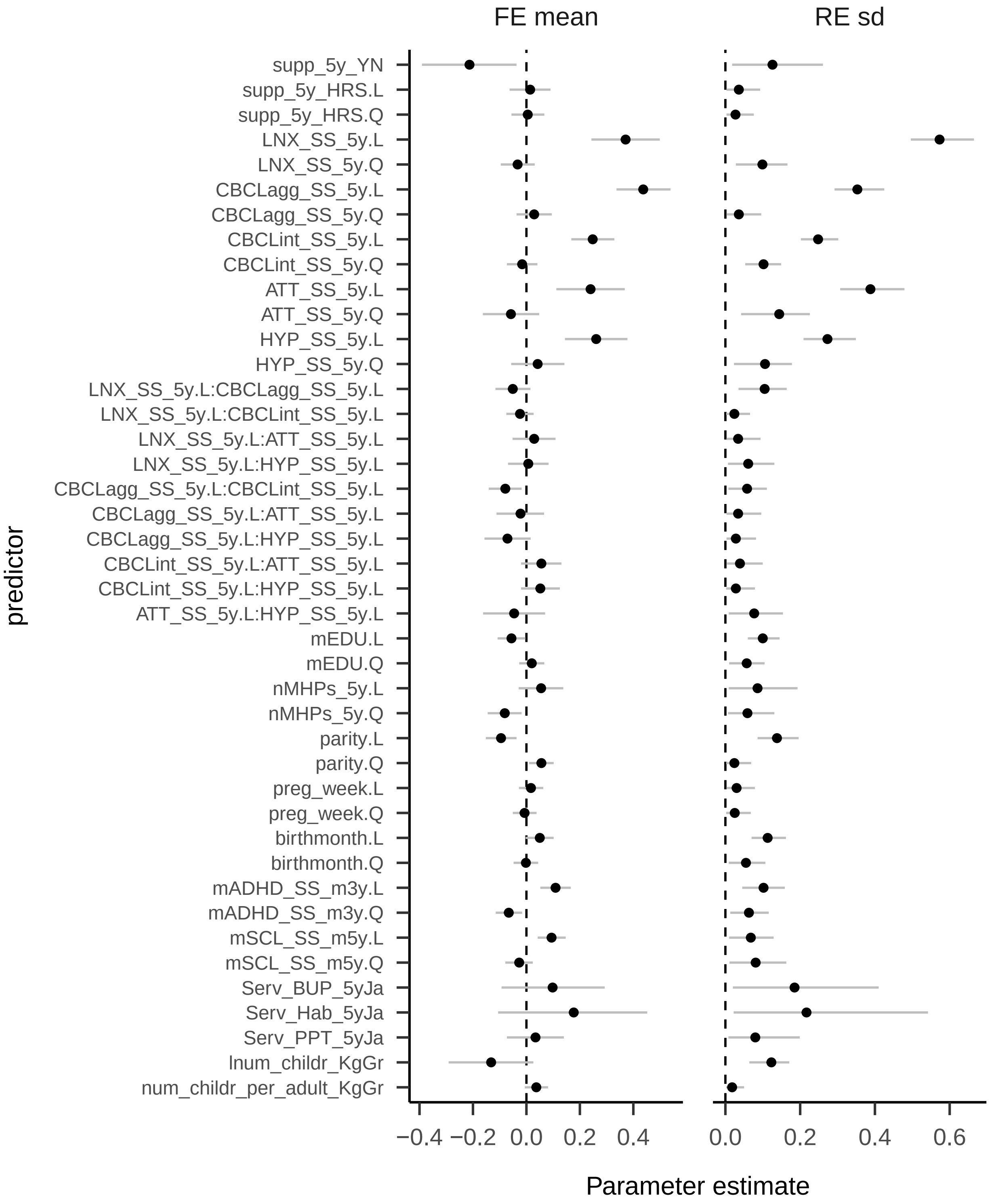


*Figure S9*. Coefficients of the hierarchical regression model with adjustment. FE mean = fixed effects, RE sd = standard deviation of random effects. L = linear effects, Q = quadratic effects. Colons between variables indicate interactions. The plot shows means as dots and 90% credible intervals as vertical lines.

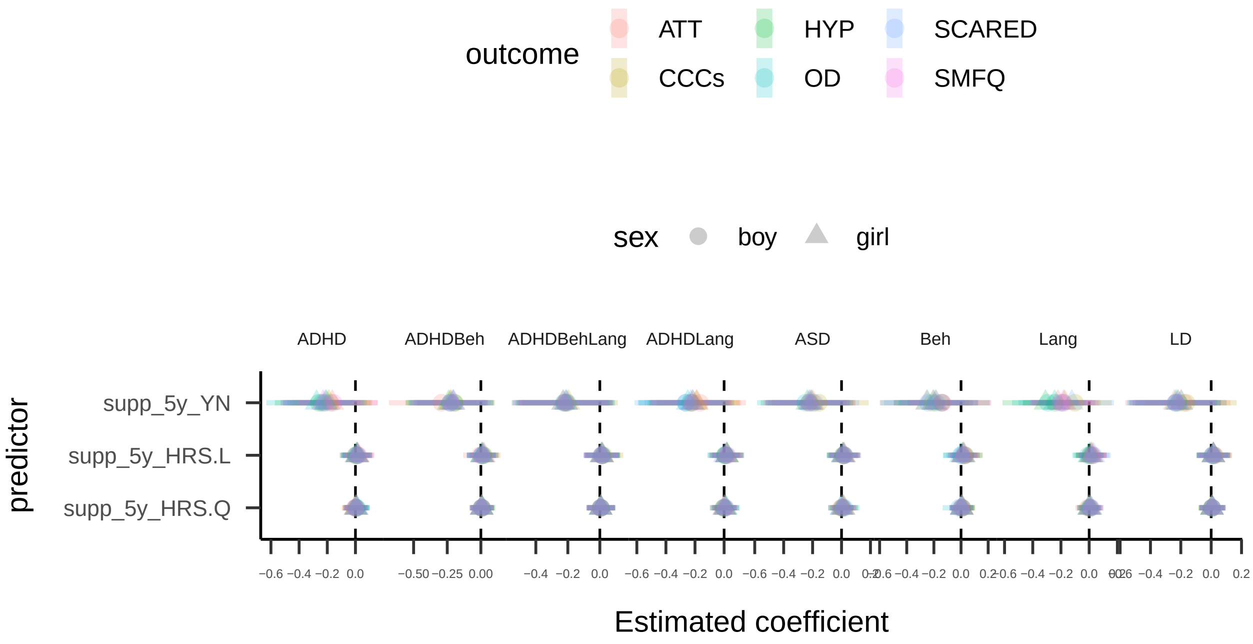


*Figure S10*. Group specific regression coefficients for the model with adjustment (fixed effects + random effects): Special educational assistance.

**Estimated average treatment effects.** Average treatment effects are only reported for the model with adjustment, because the model without adjustment produces obviously biased results (due to treatment by indication).

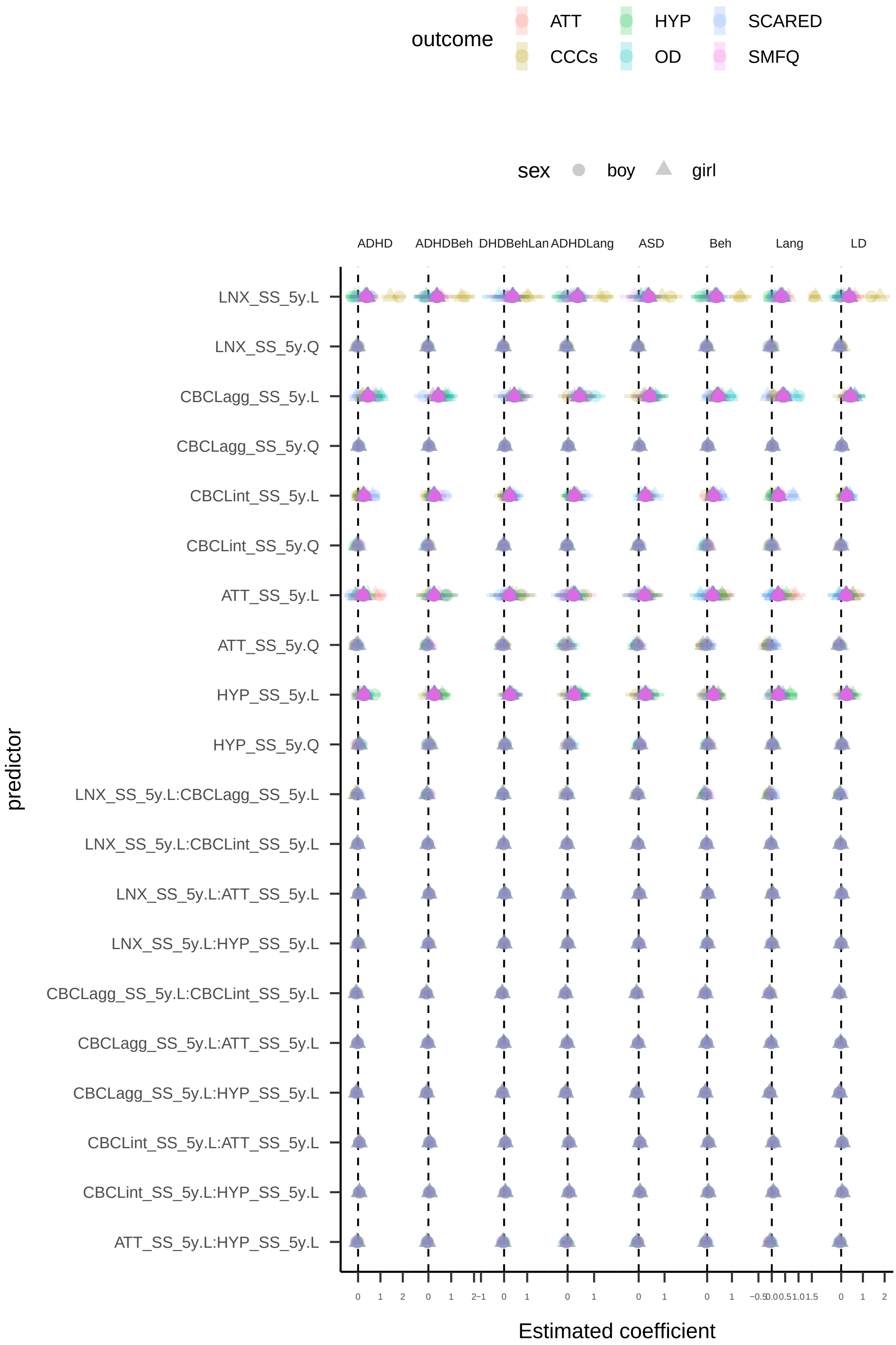


*Figure S11.* Group specific regression coefficients for the model with adjustment (fixed effects + random effects): Difficulties at baseline.

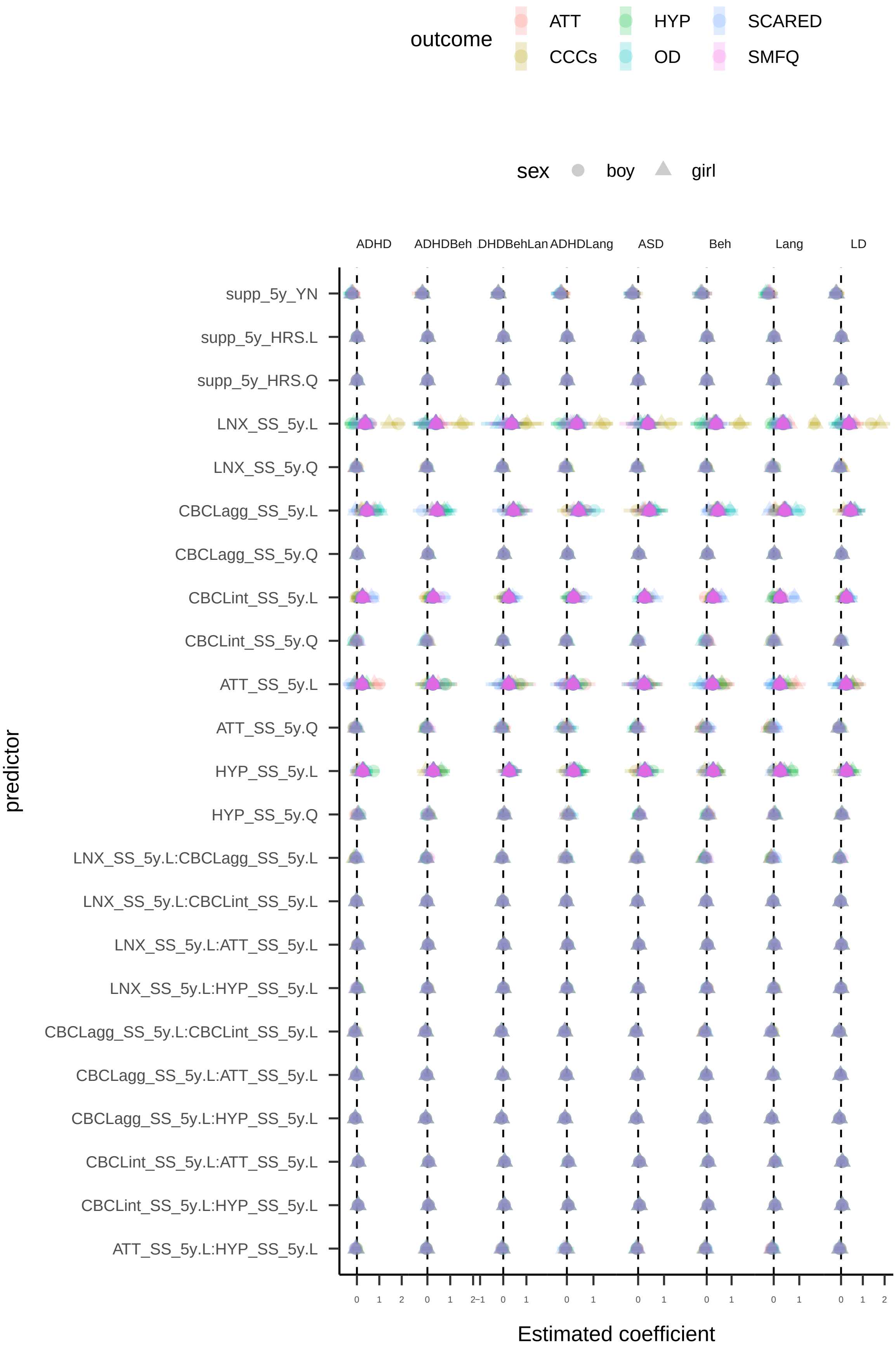


*Figure S12*. Group specific regression coefficients for the model with adjustment (fixed effects + random effects): Maternal characteristics and ECEC.

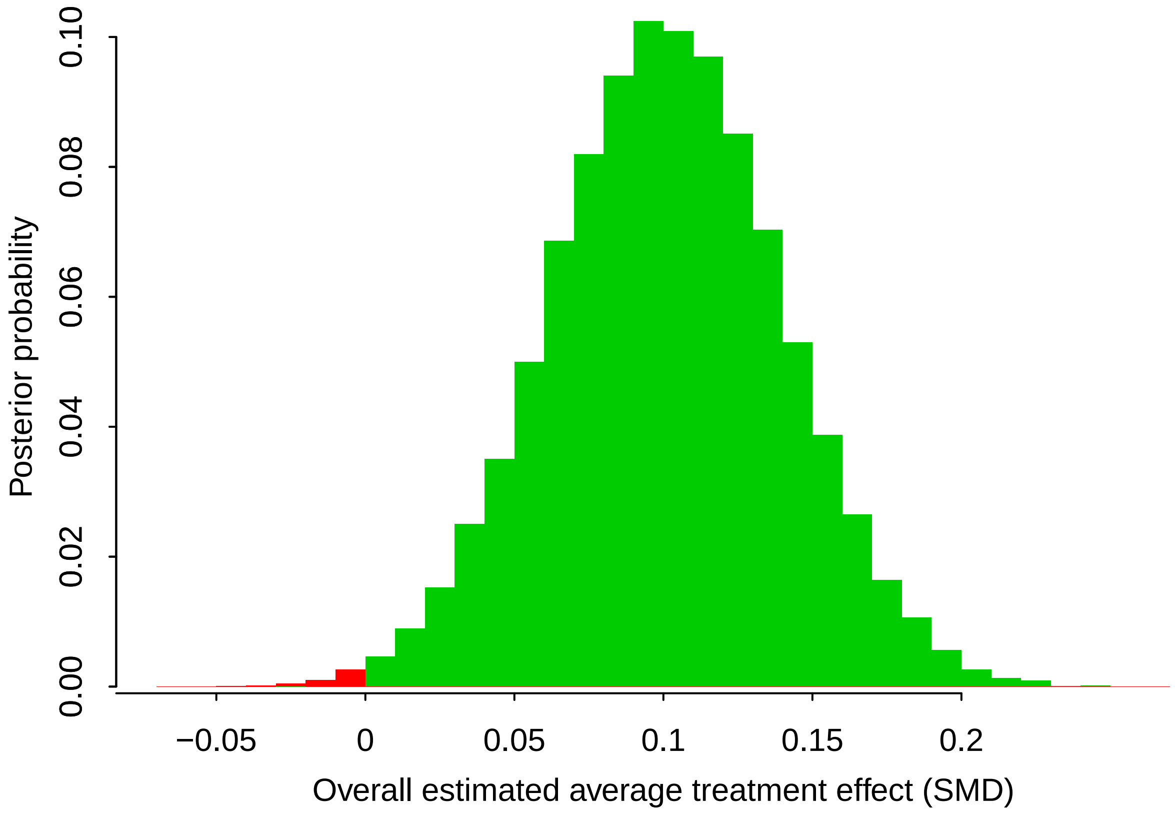


*Figure S13*. Posterior distribution of the average effect of special pedagogical assistance across all psycho-social difficulties in the study sample. The effect size is on the x-axis, the posterior probability of an effect size is on the y axis. Positive effects are displayed in green, negative effects in red. The posterior probability of a positive effect of SEA is 99.99%.

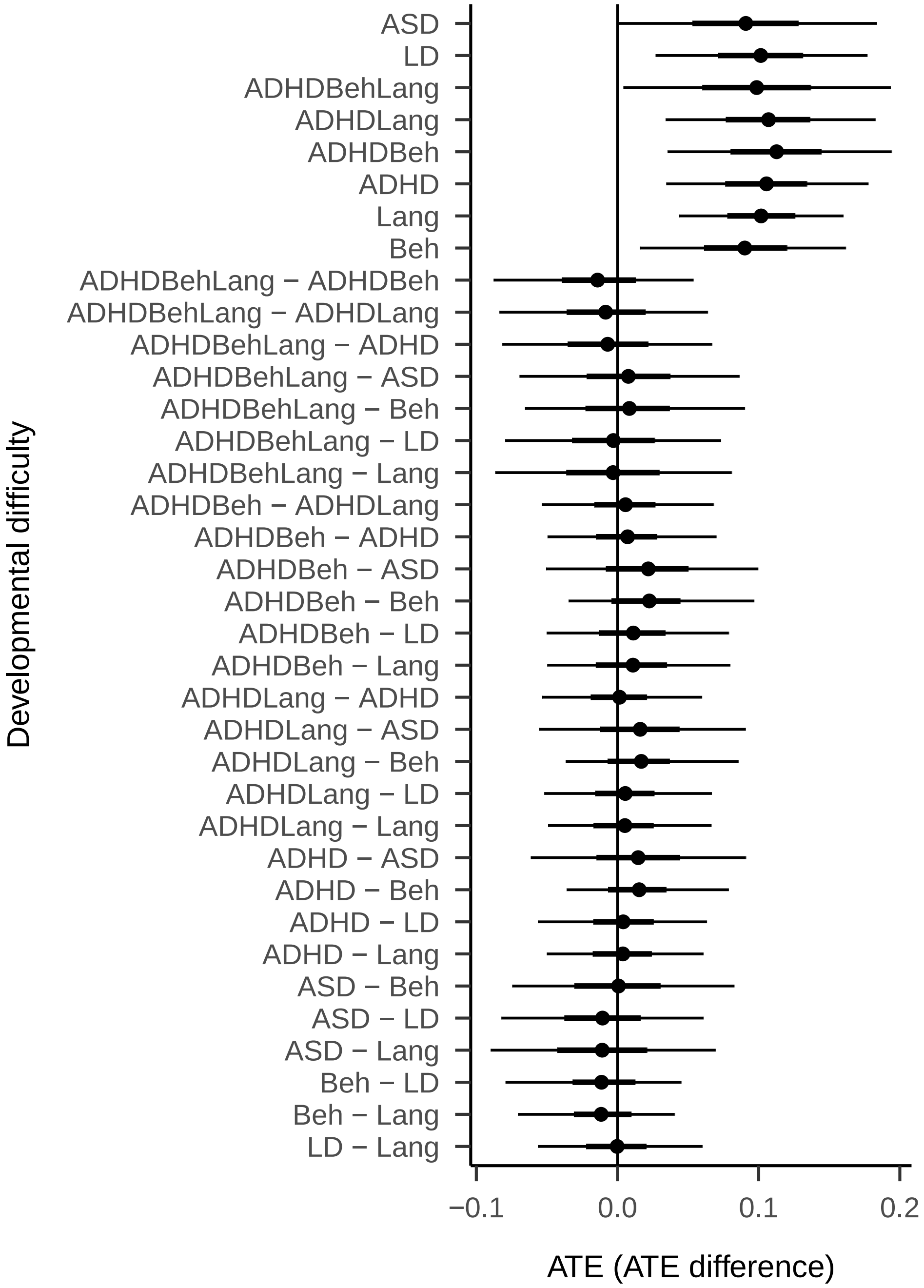


*Figure S14*. Estimated average treatment effects (ATE) and effect differences for children with different behavioral or developmental Problems. The top 6 rows show effects for the different groups (for each group averaged overall all psycho-social difficulties). The remaining rows show all pair-wise comparisons between groups. Dots are means, thick and thin lines are 50% and 90% credible intervals. Lines show 50% and 90% credible intervals.

Table S5

*SEA effects and effect differences for children with different developmental difficulties.*

| | ASD | LD | ADHDBehLang | ADHDLang | ADHDBeh | ADHD | Lang | Beh |
|---|---|---|---|---|---|---|---|---|
| ASD | .091 (0,.184) | -.011 (-.082,.061) | .008 (-.069,.086) | .016 (-.055,.091) | .022 (-.051,.099) | .015 (-.061,.091) | -.011 (-.09,.069) | .001 (-.075,.083) |
| LD | -.011 (-.082,.061) | .102 (.027,.177) | -.003 (-.08,.073) | .006 (-.052,.067) | .011 (-.05,.079) | .004 (-.056,.063) | 0 (-.056,.06) | -.011 (-.079,.045) |
| ADHDBehLang | .008 (-.069,.086) | -.003 (-.08,.073) | .099 (.004,.193) | -.008 (-.084,.064) | -.014 (-.088,.054) | -.007 (-.082,.067) | -.003 (-.087,.081) | .008 (-.065,.09) |
| ADHDLang | .016 (-.055,.091) | .006 (-.052,.067) | -.008 (-.084,.064) | .107 (.034,.183) | .006 (-.054,.068) | .001 (-.053,.06) | .005 (-.049,.066) | .017 (-.037,.086) |
| ADHDBeh | .022 (-.051,.099) | .011 (-.05,.079) | -.014 (-.088,.054) | .006 (-.054,.068) | .113 (.035,.194) | .007 (-.05,.07) | .011 (-.05,.08) | .023 (-.035,.097) |
| ADHD | .015 (-.061,.091) | .004 (-.056,.063) | -.007 (-.082,.067) | .001 (-.053,.06) | .007 (-.05,.07) | .106 (.035,.178) | .004 (-.05,.061) | .015 (-.036,.079) |
| Lang | -.011 (-.09,.069) | 0 (-.056,.06) | -.003 (-.087,.081) | .005 (-.049,.066) | .011 (-.05,.08) | .004 (-.05,.061) | .102 (.044,.16) | -.012 (-.07,.04) |
| Beh | .001 (-.075,.083) | -.011 (-.079,.045) | .008 (-.065,.09) | .017 (-.037,.086) | .023 (-.035,.097) | .015 (-.036,.079) | -.012 (-.07,.04) | .09 (.016,.162) |

*Note.* Numbers are mean ATEs (90% credible intervals). Group wise effects are on the main diagonal and pairwise comparisons on the off diagonals.

Table S6

*SEA effects and effect differences for different outcomes.*

| | ATT | HYP | OPP | MOOD | ANX | COMM |
|---|---|---|---|---|---|---|
| ATT | .096 (.024,.168) | -.015 (-.069,.033) | -.024 (-.091,.03) | 0 (-.053,.054) | -.005 (-.068,.054) | .015 (-.041,.078) |
| HYP | -.015 (-.069,.033) | .111 (.039,.184) | -.01 (-.066,.039) | .015 (-.034,.071) | .01 (-.051,.072) | .03 (-.029,.101) |
| OPP | -.024 (-.091,.03) | -.01 (-.066,.039) | .121 (.047,.198) | .025 (-.021,.083) | .02 (-.035,.082) | .04 (-.021,.117) |
| MOOD | 0 (-.053,.054) | .015 (-.034,.071) | .025 (-.021,.083) | .096 (.025,.167) | -.005 (-.061,.045) | .015 (-.041,.078) |
| ANX | -.005 (-.068,.054) | .01 (-.051,.072) | .02 (-.035,.082) | -.005 (-.061,.045) | .101 (.026,.178) | .02 (-.038,.09) |
| COMM | .015 (-.041,.078) | .03 (-.029,.101) | .04 (-.021,.117) | .015 (-.041,.078) | .02 (-.038,.09) | .081 (.004,.156) |

*Note.* Numbers are means ATEs (90% credible intervals). Outcome wise effects are on the main diagonal and pairwise comparisons on the off diagonals.

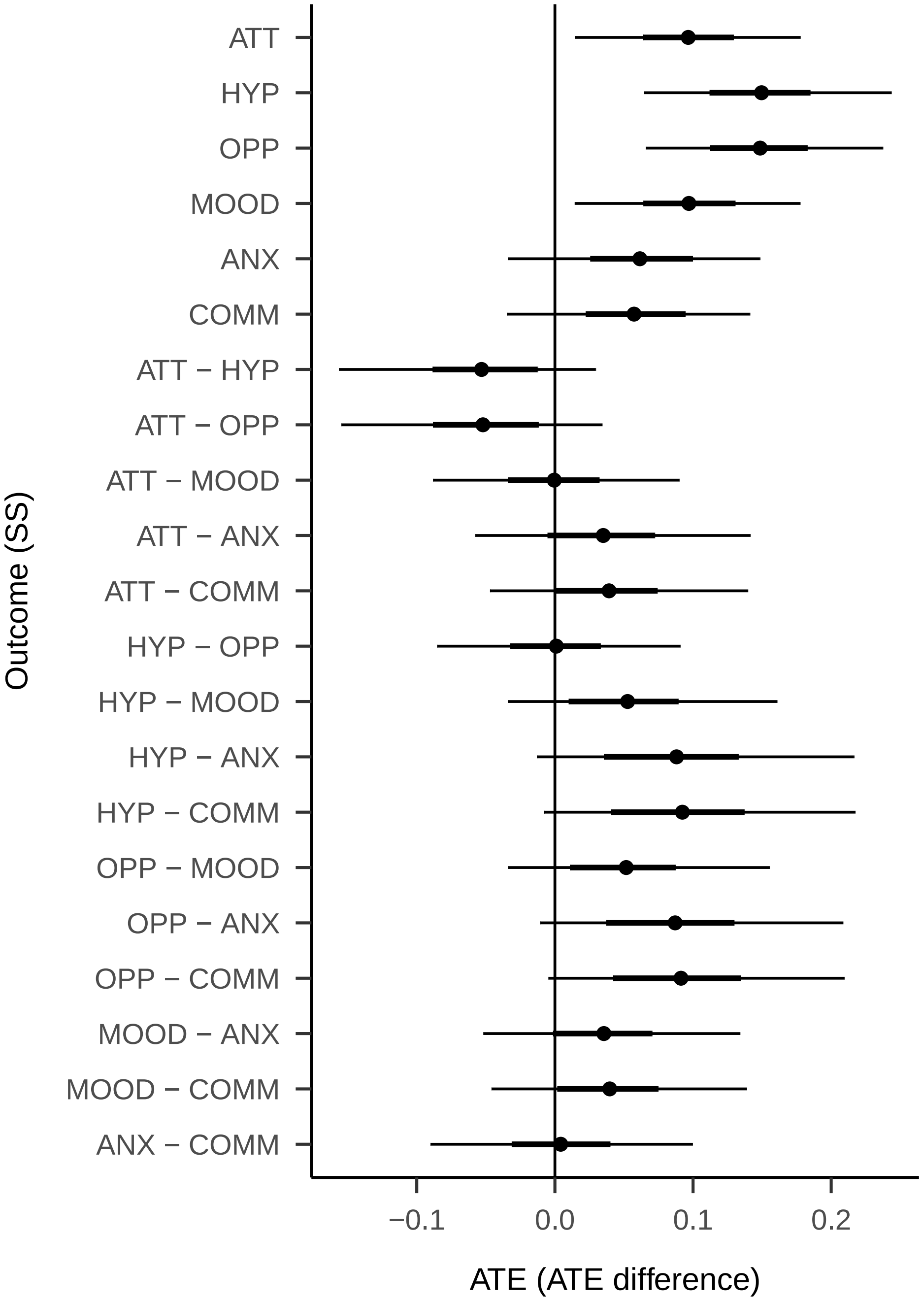


*Figure S15*. Estimated average treatment effects (ATE) and effect differences for different psycho-social difficulties. The top 6 rows show effects for the different psycho-social difficulties (for each difficulty averaged overall all DBPs). The remaining rows show all pair-wise comparisons between difficulties. Dots are means, thick and thin lines are 50% and 90% credible intervals.

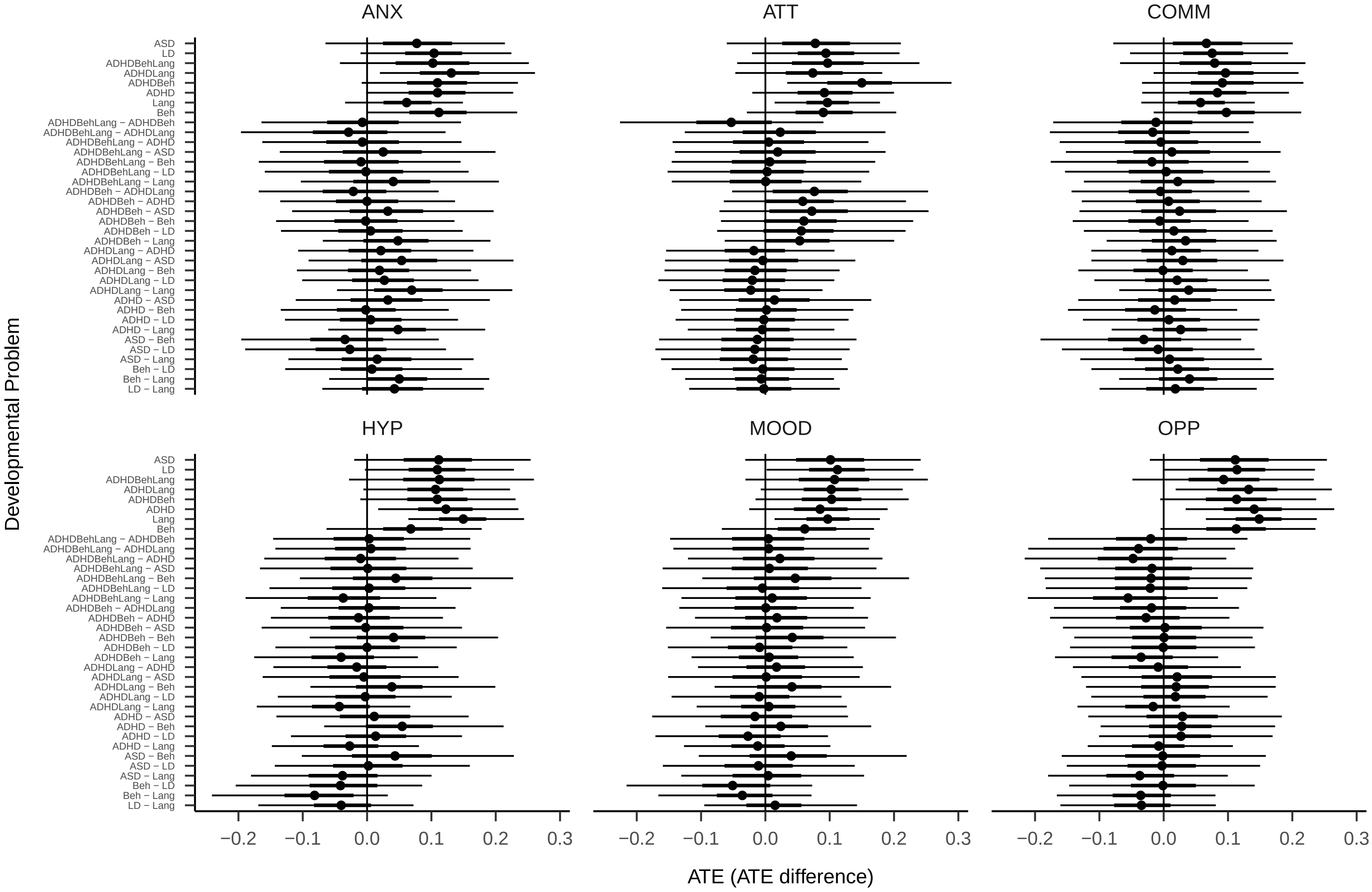


*Figure S16*. Estimated average treatment effects (ATE) and effect differences by group and outcome.

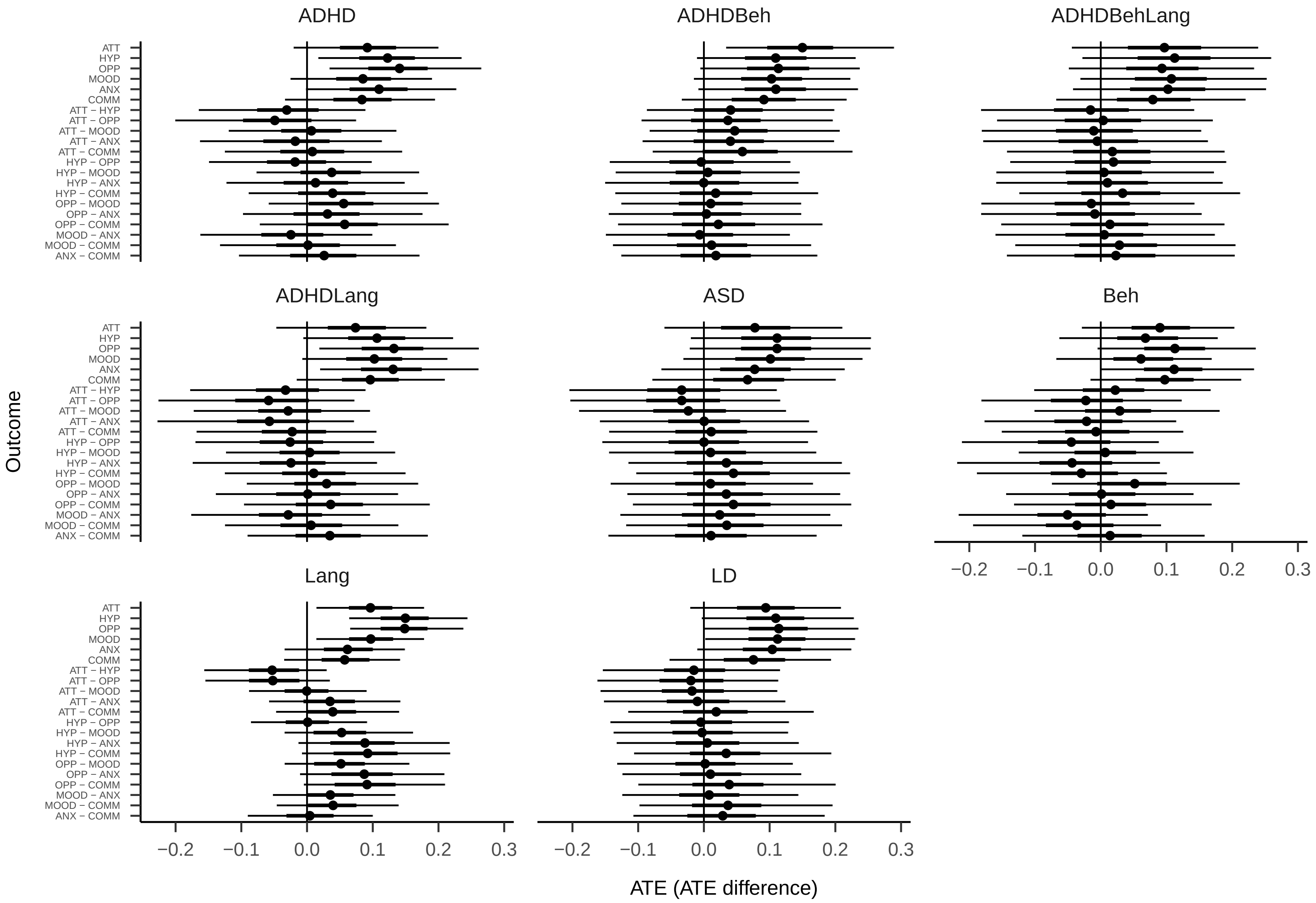


*Figure S17*. Estimated average treatment effects (ATE) and effect differences by outcome and group.

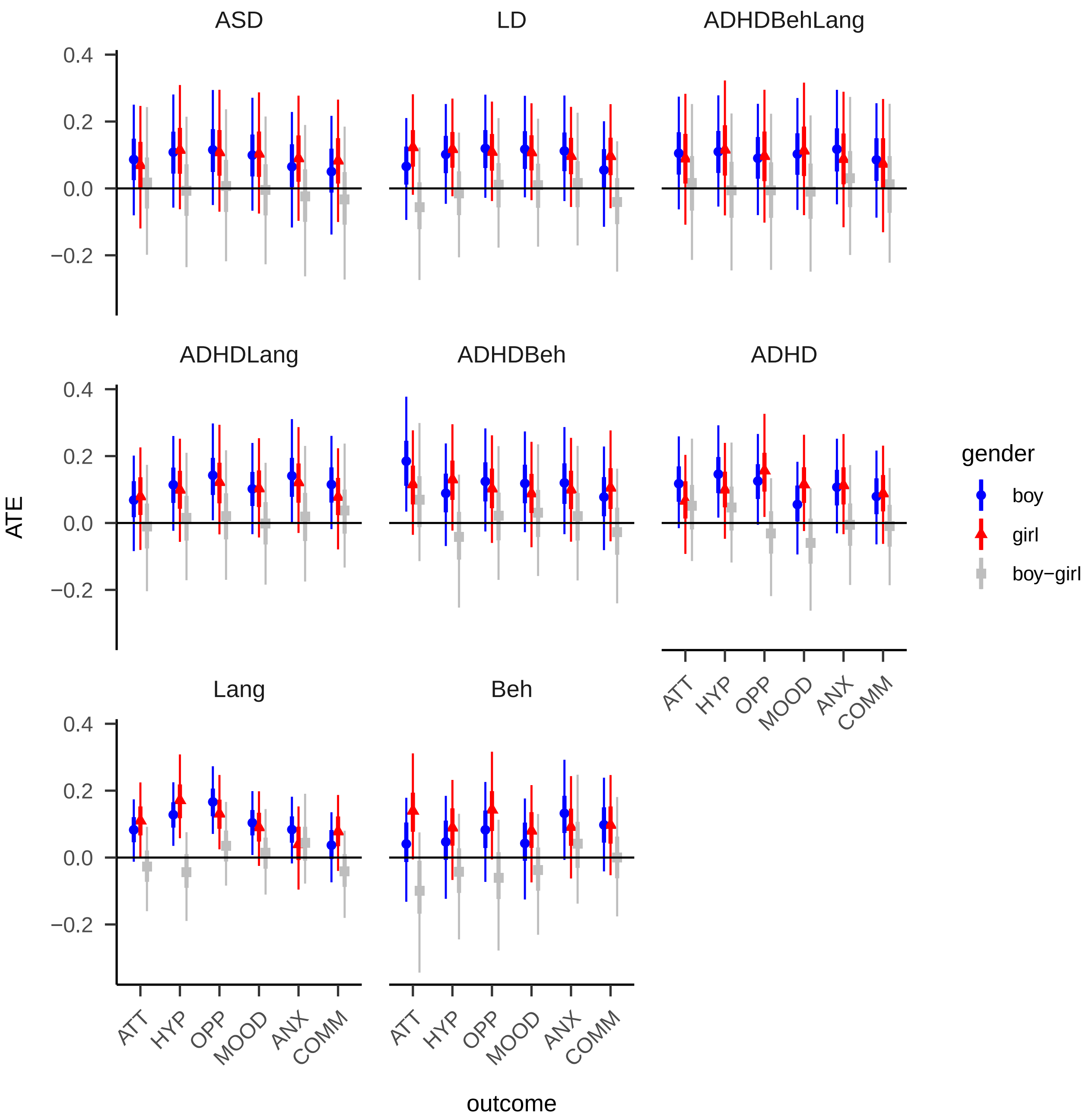


*Figure S18*. Estimated average treatment effects, split by developmental or behavioral problem, psycho-social difficulty, and gender. Dots are means, thick and thin lines are 50% and 90% credible intervals.

**Supplementary References**

1.

R Core Team (2017) R: A language and environment for statistical computing. R Foundation for Statistical Computing, Vienna, Austria

2.

Stanley D (2018) apaTables: Create american psychological association (APA) style tables

3.

Heinzen E, Sinnwell J, Atkinson E, et al (2018) Arsenal: An arsenal of 'r' functions for large-scale statistical summaries

4.

Gabry J, Mahr T (2018) Bayesplot: Plotting for bayesian models

5.

Bürkner P-C (2017) brms: An R package for Bayesian multilevel models using Stan. Journal of Statistical Software 80:1–28. https://doi.org/10.18637/jss.v080.i01

6.

Bürkner P-C (2018) Advanced Bayesian multilevel modeling with the R package brms. The R Journal 10:395–411. https://doi.org/10.32614/RJ-2018-017

7.

Zeileis A, Hornik K, Murrell P (2009) Escaping RGBland: Selecting colors for statistical graphics. Computational Statistics & Data Analysis 53:3259–3270. https://doi.org/10.1016/j.csda.2008.11.033

8.

Stauffer R, Mayr GJ, Dabernig M, Zeileis A (2009) Somewhere over the rainbow: How to make effective use of colors in meteorological visualizations. Bulletin of the American Meteorological Society 96:203–216. https://doi.org/10.1175/BAMS-D-13-00155.1

9.

Dowle M, Srinivasan A (2018) Data.table: Extension of 'data.frame'

10.

al. AS et mult. (2019) DescTools: Tools for descriptive statistics

11.

Fox J, Venables B, Damico A, Salverda AP (2019) English: Translate integers into english

12.

Gohel D (2020) Flextable: Functions for tabular reporting

13.

Zeileis A, Croissant Y (2010) Extended model formulas in R: Multiple parts and multiple responses. Journal of Statistical Software 34:1–13. https://doi.org/10.18637/jss.v034.i01

14.

Wickham H (2016) ggplot2: Elegant graphics for data analysis. Springer-Verlag New York

15.

Kassambara A (2019) Ggpubr: ’ggplot2’ based publication ready plots

16.

Gordon M (2018) Gmisc: Descriptive statistics, transition plots, and more

17.

Harrell Jr FE, Charles Dupont with contributions from, others. many (2018) Hmisc: Harrell miscellaneous

18.

Gordon M, Gragg S, Konings P (2018) htmlTable: Advanced tables for markdown/HTML

19.

Long JA (2019) Jtools: Analysis and presentation of social scientific data

20.

Zhu H (2019) kableExtra: Construct complex table with ’kable’ and pipe syntax

21.

Xie Y (2015) Dynamic documents with R and knitr, 2nd ed. Chapman; Hall/CRC, Boca Raton, Florida

22.

Sarkar D (2008) Lattice: Multivariate data visualization with r. Springer, New York

23.

Bache SM, Wickham H (2014) Magrittr: A forward-pipe operator for r

24.

Gohel D (2021) Officer: Manipulation of microsoft word and PowerPoint documents

25.

Aust F, Barth M (2018) papaja: Create APA manuscripts with R Markdown

26.

Eddelbuettel D, François R (2011) Rcpp: Seamless R and C++ integration. Journal of Statistical Software 40:1–18. https://doi.org/10.18637/jss.v040.i08

27.

Eddelbuettel D, Balamuta JJ (2017) Extending extitR with extitC++: A Brief Introduction to extitRcpp. PeerJ Preprints 5:e3188v1. https://doi.org/10.7287/peerj.preprints.3188v1

28.

Stan Development Team (2018) RStan: The R interface to Stan

29.

Goodrich B, Gabry J, Ali I, Brilleman S (2018) Rstanarm: Bayesian applied regression modeling via Stan.

30.

Ooms J, Hester J (2019) Spelling: Tools for spell checking in r

31.

Stan Development Team (2018) StanHeaders: Headers for the R interface to Stan

32.

Terry M. Therneau, Patricia M. Grambsch (2000) Modeling survival data: Extending the Cox model. Springer, New York

33.

Buuren S van, Groothuis-Oudshoorn K (2011) Mice: Multivariate imputation by chained equations in r. Journal of Statistical Software 45:1–67

34.

Goodrich B, Gabry J, Ali I, Brilleman S (2019) rstanarm: Bayesian applied regression modeling via Stan

35.

Goodrich B, Gabry J, Ali I, Brilleman S (2018) Rstanarm: Bayesian applied regression modeling via Stan.

36.

Austin PC, Stuart EA (2015) Moving towards best practice when using inverse probability of treatment weighting (IPTW) using the propensity score to estimate causal treatment effects in observational studies. Statistics in medicine 34:3661–3679

37.

Bürkner P-C (2017) brms: An R package for Bayesian multilevel models using Stan. Journal of Statistical Software 80:1–28. https://doi.org/10.18637/jss.v080.i01

38.

Bürkner P-C (2018) Advanced Bayesian multilevel modeling with the R package brms. The R Journal 10:395–411. https://doi.org/10.32614/RJ-2018-017

39.

Gelman A, Rubin DB (1992) Inference from iterative simulation using multiple sequences. Statistical science: a review journal of the Institute of Mathematical Statistics 7:457–472

40.

Root A, Brown JP, Forbes HJ, et al (2019) Association of Relative Age in the School Year With Diagnosis of Intellectual Disability, Attention-Deficit/Hyperactivity Disorder, and Depression. JAMA pediatrics
